# Genomic and clinical determinants of extraintestinal *Clostridium perfringens* infections in immunocompromised patients

**DOI:** 10.64898/2026.02.18.26346578

**Authors:** Basma Menif, Samantha E. Wirth, Danielle Wroblewski, Julia A. Connors, Nidia Correa, Mary Louise Delaney, Lynn Bry

## Abstract

**Background:** *Clostridium perfringens* can cause life-threatening extraintestinal infections in immunocompromised patients, an area in which we have little information regarding strain factors that impact patient risks and outcomes.

**Methods:** We conducted genomic-epidemiologic analyses on *C. perfringens* isolates from 70 patients seen at Brigham and Women’s Hospital over 2021–2024. Genomic analyses evaluated strain profiles within a broader context of 2,321 *C. perfringens* genomes from foodborne, veterinary, clinical, and environmental sources to identify factors associated with invasive infections.

**Results:** Of 70 patients with *C. perfringens* infections (mean age 67.6 years), 32 had invasive infections, of which two-thirds had active malignancies, and more than half were immunocompromised. Patients with invasive infections had a significantly higher 90-day mortality of 43.8% vs. 18.4% (p=0.035) and a higher median Charlson Comorbidity Index (6 vs. 3; p=0.003). Notably, no patient isolates were clonal, verifying the absence of hospital-based transmission. Patient isolates showed increased carriage of hyaluronidases (*nagHIJKL*), sialidases (*nanIJ*), and perfringolysin O (*pfoA*). Genomic-epidemiologic analyses identified a new independent association between the NagL hyaluronidase (OR 3.90, 95% CI 1.14 - 16.24) in highly morbid invasive infections.

**Conclusion:** We present a comprehensive genomic analysis of *C. perfringens* and of strains infecting immunocompromised patients, including epidemiologic associations of the hyluronidase NagL, NanIJ sialidase, and perfringolysin O in highly morbid invasive infections. These genes provide potential markers to identify high morbidity strains that can infect these populations and to further elucidate their role in invasive infections.

## Introduction

*Clostridium perfringens* is one of the most common intestinal pathogens in humans and animals, causing nearly one million cases of foodborne diseases in the United States and resulting in substantial economic losses in the animal agriculture industry [1]. *C. perfringens* can also cause extra-intestinal infections, from seeding after blunt trauma or after hepatobiliary or gastrointestinal procedures, leading to sepsis as well as necrotizing skin and soft tissue infections, which have high morbidity and mortality, especially in immunocompromised patients [2–4]. Although uncommon, invasive infections occur most frequently in individuals with hepatobiliary, pancreatic, or hematologic malignancies, and with mortality rates ranging from 27-44% [3–6]. Anti-neoplastic therapies that disrupt the intestinal mucosal barrier are thought to facilitate bacterial translocation from the gut, with dissemination further impacted by a patient’s immune status. However, whether strains causing invasive infections have distinct virulence features compared with strains associated with colonization or foodborne disease has not been well defined, but opens opportunities to identify virulence factors and means to better informed prevention and treatment of these infections in highly vulnerable patient populations.

*C. perfringens* is a ubiquitous and anaerobic spore-forming bacterium that frequently colonizes the mammalian gut. Pathogenic strains produce diverse extracellular toxins as well as enzymes that support its metabolism but can also rapidly damage host tissues [2]. *C. perfringens* isolates are categorized into seven toxinotypes (A to G) based on the presence of six major toxins (α, β, ɛ, ɩ, enterotoxin, and *netB*), each associated with distinct host specificities and specific diseases [7]. Toxinotype A strains, with the toxin *cpa* gene, are associated with enteric and histotoxic infections. Toxinotype F strains produce enterotoxin (CPE) and occur primarily in human food poisoning, whereas other toxinotypes primarily cause infections in animals, particularly poultry (G), and ruminants and pigs (B-E) [7,8]. Beyond typing toxins, *C. perfringens* produces additional toxins and enzymes, such as perfringolysin O, sialidases, and hyaluronidases, that contribute to tissue destruction and disease progression [8,9].

Prior genomic investigations demonstrated five phylogroups (I-V) that possessed distinct disease features and host specificities [9,10], particularly of foodborne outbreaks or necrotic enteritis in poultry [11,12]. These studies highlighted the role of plasmid-encoded toxins, horizontal gene transfer, and accessory genes in *C. perfringens’* pathogenicity. However, genomic investigations of *C. perfringens* causing extra-intestinal infections in humans are lacking despite their clinical significance, especially in immunocompromised persons who are at elevated risk for life-threatening invasive infections [3].

We analyzed extra-intestinal *C. perfringens* infections in highly immunocompromised patients seen at Brigham and Women’s Hospital from 2021-2024. Strain genomic analyses evaluated pathogen genetic profiles within the context of globally deposited *C. perfringens* genomes indexed in the National Center for Biotechnology Information (NCBI) Pathogen Detection program [13,14] to evaluate strain population structures and virulence determinants associated with invasive infections.

## Methods

### IRB study protocol and data collection

We conducted a retrospective clinical and genomic analysis on 78 *C. perfringens* clinical strains isolated from 74 BWH patients from 2021-2024. The study was conducted under IRB protocol 2011-P-002883 (PI: Bry, Mass-General-Brigham). We used the Crimson LIMS to retrieve *C. perfringens* positive samples [15]. Patient data and Labs, including microbiological test results, were searched using the Partners Research Patient Data Registry [16]. The Charlson Comorbidity Index was used to assess comorbidities, and clinical severity was evaluated with the Sequential Organ Failure Assessment (SOFA) score. Invasive infections were defined as blood systemic infections, deep abscesses, and necrotizing soft tissue infections.

Comparative genomic analyses evaluated publicly available *C. perfringens* genomes from the NCBI Pathogen Detection as of June 2025 (*n*= 2,570) with associated metadata on country, year, and source of isolation [17].

### Whole-genome sequencing

Whole-genome sequencing (WGS) was performed on the Illumina NextSeq500 platform at the Wadsworth Center Bacteriology Laboratory, New York State Department of Health, following a previously published protocol [18]. Sequence data was further assessed for quality using MicroRunQC on GalaxyTrakr[13]. Sequences were manually uploaded to the NCBI Sequence Read Archive (SRA) using the One Health Enteric Metadata template. Strain international placement was performed using SNP clustering in NCBI Pathogen Detection [17].

### Phylogenetic analysis

Analyses investigated 2,321 high-quality *C. perfringens* genome assemblies that passed contamination checks via CheckM v1.2.3 (completeness >80% and contamination <5%). *C. perfringens* genome assemblies were dereplicated at the strain level (ANI > 99%) using dRep v.3.6.2 [19], to provide a non-redundant dataset of 441 representative genomes, including all BWH isolates and representative strains from each phylogroup.

Coding sequences were annotated with Prokka v.1.14.6, and a core-gene alignment, comprising 1874 single-copy core genes, was subsequently constructed using Panaroo v.1.5.2 [20]. SNPs were next extracted from core-gene alignments using snp-sites v.2.5.1 [21] and maximum-likelihood trees were constructed using IQ-TREE v.2.2.6 [22] with general time-reversible (GTR)-gamma model and ultra-fast 1000 bootstrap replicates. Tree annotation was performed via iTOL v.7. Phylogroups were defined based on monophyletic clustering in the core-genome phylogeny, supported by pairwise ANI values. The tree is available online at https://itol.embl.de/tree/702214621778001758204596 MLST was determined *in silico* using the MLST tool with the *C. perfringens* PubMLST scheme.

### In *silico* analysis of virulence-related genes

Virulence genes were identified using ABRicate v.1.0.1 (https://github.com/tseemann/abricate) with a 90% identity and 60% coverage threshold, based on a custom sequence database that included 76 up-to-date virulence factors reported for *C. perfringe*ns [9]. Toxinotypes A-G were assigned via Abricate and Pathogen detection database [7].

### Statistical analysis

Categorical data were compared using the chi-squared or Fisher’s exact test, and continuous data using a t-test and ANOVA or the Mann-Whitney and Kruskal-Wallis tests, as appropriate. P values <0.05 were considered statistically significant after Benjamini-Hochberg correction for multiple comparisons.

To validate candidate virulence genes enriched in human extra-intestinal isolates and to minimize false positive association caused by strain clonality, especially among intestinal isolates, we performed a pan-genome-wide association study (pan-GWAS) using PySeer accounting for population structure (v1.3.9) [23]. We restricted analyses to genes with MAF >0.25, providing >80% power to detect association with odds ratios (OR) >2 in our dataset of 920 human genomes (PowerBacGWAS, Supplementary Figure S1) [24]. Gene presence/absence matrices were derived from the Panaroo output, and population structure was controlled using the first 20 multidimensional scaling (MDS) components from the pairwise genetic distance matrix. Associations were tested by fixed-effects logistic regression with Benjamini-Hochberg correction.

Univariate and multivariate models were developed to assess virulence-related genes associated with *C. perfringens* invasive infections, adjusting for clinically relevant covariates. Given the large number of determinants and small sample size, associations with invasiveness (binary outcome) were tested using penalized logistic regression with Firth’s correction, adjusting for age, sex, Charlson Comorbidity Index, immunosuppression, and polymicrobial infection. Candidate genes were selected using penalized regression with the least absolute shrinkage and selection operator (LASSO) implemented in the glmnet R package, with leave-one-out cross-validation to tune the penalty parameter. Regression analyses were restricted to 70 isolates with complete data for all variables. With 32 invasive and 38 non-invasive isolates, the study had >60% power to detect associations with OR ≥ 3 for genes with intermediate prevalence (30–60%), based on two-proportion tests using Cohen’s h (α = 0.05).

All analyses were performed in R version 4.4.2 (www.R-project.org). The study was reported according to STROME-ID guidelines (Supplementary Table S1)

## Results

### BWH *C. perfringens* isolates are predominantly associated with invasive infections

From 2021-2024, 74 patients developed clinically diagnosed infections with *C. perfringens,* resulting in 78 clinical strains (Table 1). Complete clinical data was available for 70 patients, with a mean age of 67.6 ± 16.3 years.

**Table 1:**
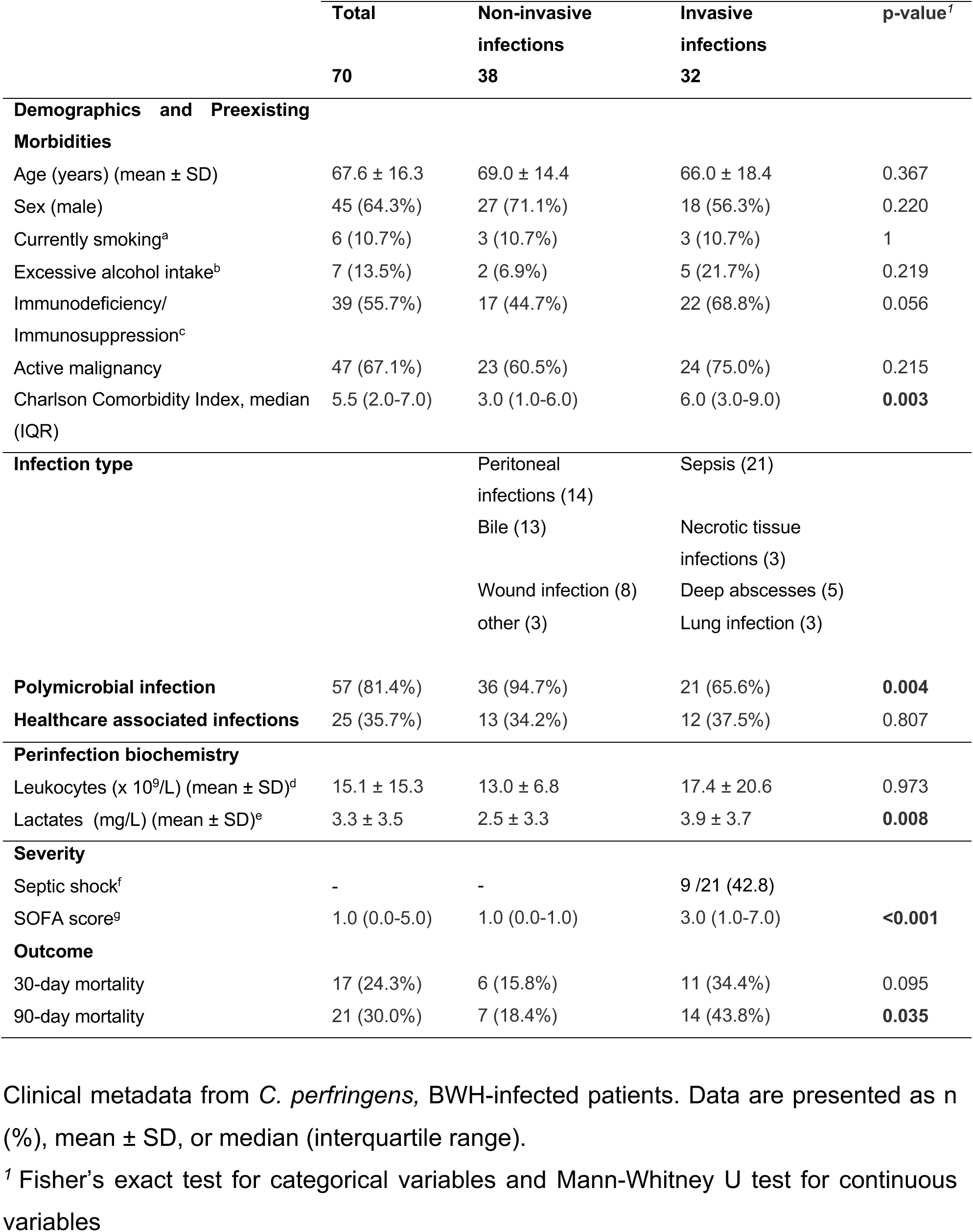
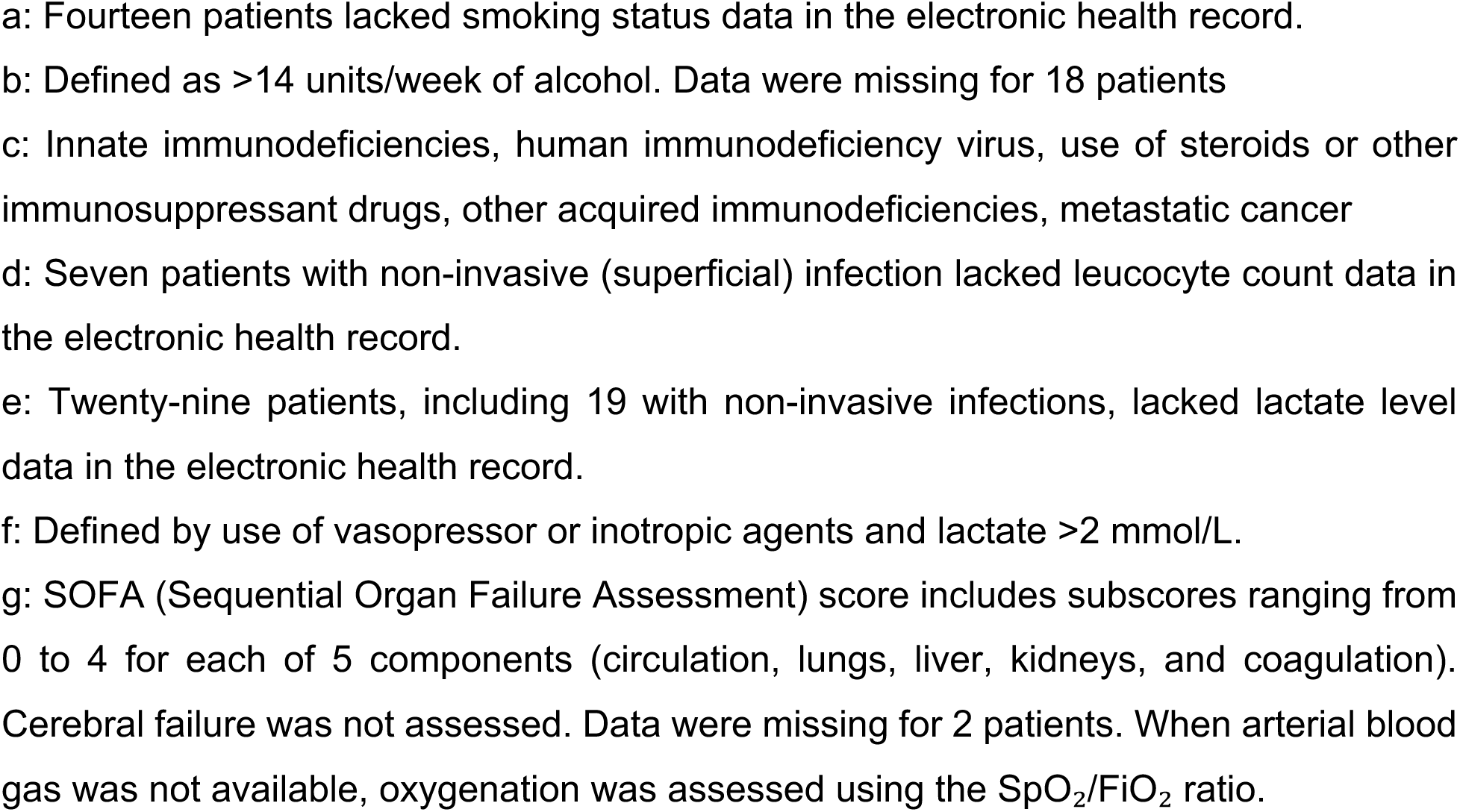
Clinical characteristics of *C. perfringens-*infected patients.

|  | Total<br>70 | Non-invasive<br>infections<br>38 | Invasive<br>infections<br>32 | p-value<sup>1</sup> |
| --- | --- | --- | --- | --- |
| **Demographics and Preexisting Morbidities** |  |  |  |  |
| Age (years) (mean ± SD) | 67.6 ± 16.3 | 69.0 ± 14.4 | 66.0 ± 18.4 | 0.367 |
| Sex (male) | 45 (64.3%) | 27 (71.1%) | 18 (56.3%) | 0.220 |
| Currently smoking<sup>a</sup> | 6 (10.7%) | 3 (10.7%) | 3 (10.7%) | 1 |
| Excessive alcohol intake<sup>b</sup> | 7 (13.5%) | 2 (6.9%) | 5 (21.7%) | 0.219 |
| Immunodeficiency/<br>Immunosuppression<sup>c</sup> | 39 (55.7%) | 17 (44.7%) | 22 (68.8%) | 0.056 |
| Active malignancy | 47 (67.1%) | 23 (60.5%) | 24 (75.0%) | 0.215 |
| Charlson Comorbidity Index, median<br>(IQR) | 5.5 (2.0-7.0) | 3.0 (1.0-6.0) | 6.0 (3.0-9.0) | **0.003** |
| **Infection type** |  |  |  |  |
|  |  | Peritoneal<br>infections (14) | Sepsis (21) |  |
|  |  | Bile (13) | Necrotic tissue<br>infections (3) |  |
|  |  | Wound infection (8) | Deep abscesses (5) |  |
|  |  | other (3) | Lung infection (3) |  |
| **Polymicrobial infection** | 57 (81.4%) | 36 (94.7%) | 21 (65.6%) | **0.004** |
| **Healthcare associated infections** | 25 (35.7%) | 13 (34.2%) | 12 (37.5%) | 0.807 |
| **Perinfection biochemistry** |  |  |  |  |
| Leukocytes (x 10<sup>9</sup>/L) (mean ± SD)<sup>d</sup> | 15.1 ± 15.3 | 13.0 ± 6.8 | 17.4 ± 20.6 | 0.973 |
| Lactates (mg/L) (mean ± SD)<sup>e</sup> | 3.3 ± 3.5 | 2.5 ± 3.3 | 3.9 ± 3.7 | **0.008** |
| **Severity** |  |  |  |  |
| Septic shock<sup>f</sup> | - | - | 9 /21 (42.8) |  |
| SOFA score<sup>g</sup> | 1.0 (0.0-5.0) | 1.0 (0.0-1.0) | 3.0 (1.0-7.0) | **<0.001** |
| **Outcome** |  |  |  |  |
| 30-day mortality | 17 (24.3%) | 6 (15.8%) | 11 (34.4%) | 0.095 |
| 90-day mortality | 21 (30.0%) | 7 (18.4%) | 14 (43.8%) | **0.035** |

Clinical metadata from *C. perfringens*, BWH-infected patients. Data are presented as n (%), mean ± SD, or median (interquartile range). <sup>1</sup> Fisher's exact test for categorical variables and Mann-Whitney U test for continuous variables
a: Fourteen patients lacked smoking status data in the electronic health record. b: Defined as >14 units/week of alcohol. Data were missing for 18 patients c: Innate immunodeficiencies, human immunodeficiency virus, use of steroids or other immunosuppressant drugs, other acquired immunodeficiencies, metastatic cancer d: Seven patients with non-invasive (superficial) infection lacked leucocyte count data in the electronic health record. e: Twenty-nine patients, including 19 with non-invasive infections, lacked lactate level data in the electronic health record. f: Defined by use of vasopressor or inotropic agents and lactate >2 mmol/L. g: SOFA (Sequential Organ Failure Assessment) score includes subscores ranging from 0 to 4 for each of 5 components (circulation, lungs, liver, kidneys, and coagulation). Cerebral failure was not assessed. Data were missing for 2 patients. When arterial blood gas was not available, oxygenation was assessed using the SpO<sub>2</sub>/FiO<sub>2</sub> ratio.

Thirty-day and 90-day mortality were high, at 24.3% and 30.0%, respectively. Concomitantly, the infected cohort had a high comorbidity Charlson index of 5.3, with 67% of patients having active malignancies and more than half being immunocompromised (Table 1). Invasive infections occurred in 32 patients, with sepsis occurring in 21 cases, 10 of which were mono-microbial and 2 of which were complicated by severe intravascular hemolysis. Among the invasive tissue infections, 66 % were polymicrobial. Invasive infections were associated with greater comorbidity, higher severity, and increased 90-day mortality (Table 1). In contrast, non-invasive infections were predominantly polymicrobial (95%) and largely associated with peritonitis or infections of the hepatobiliary tract (Table 1).

### BWH *C. perfringens* were non-clonal but clustered primarily within phylogroup III

To examine the epidemiological and phylogenetic relationships of BWH isolates and identify potential sources or reservoirs, we evaluated their genomic findings with strain genomes deposited in NCBI Pathogen detection from other global populations [13]. Phylogenetic analyses of globally deposited *C. perfringens* showed the five known distinct phylogroups and host-associations (Figure 1) [9,10]. Phylogroup I (n=202 strains) was associated with human foodborne diseases (n=168, 83%, *cpe*^+^) (Table 2). Phylogroup II (n=106) included human and animal isolates from toxinotypes A, E, and *netF*-positive toxinotype F (Supplementary Figure S2, Table S2). Phylogroup III (n = 1,912 strains) was the largest and most diverse group, comprising strains from different hosts and diseases (Table 2). Phylogroups IV (n=44) and V (n=56) contained only toxinotype A strains. MLST assigned 47% of the 2,321 strains to 340 STs, most of which were rare (Supplementary Table S3).

**Figure 1:**
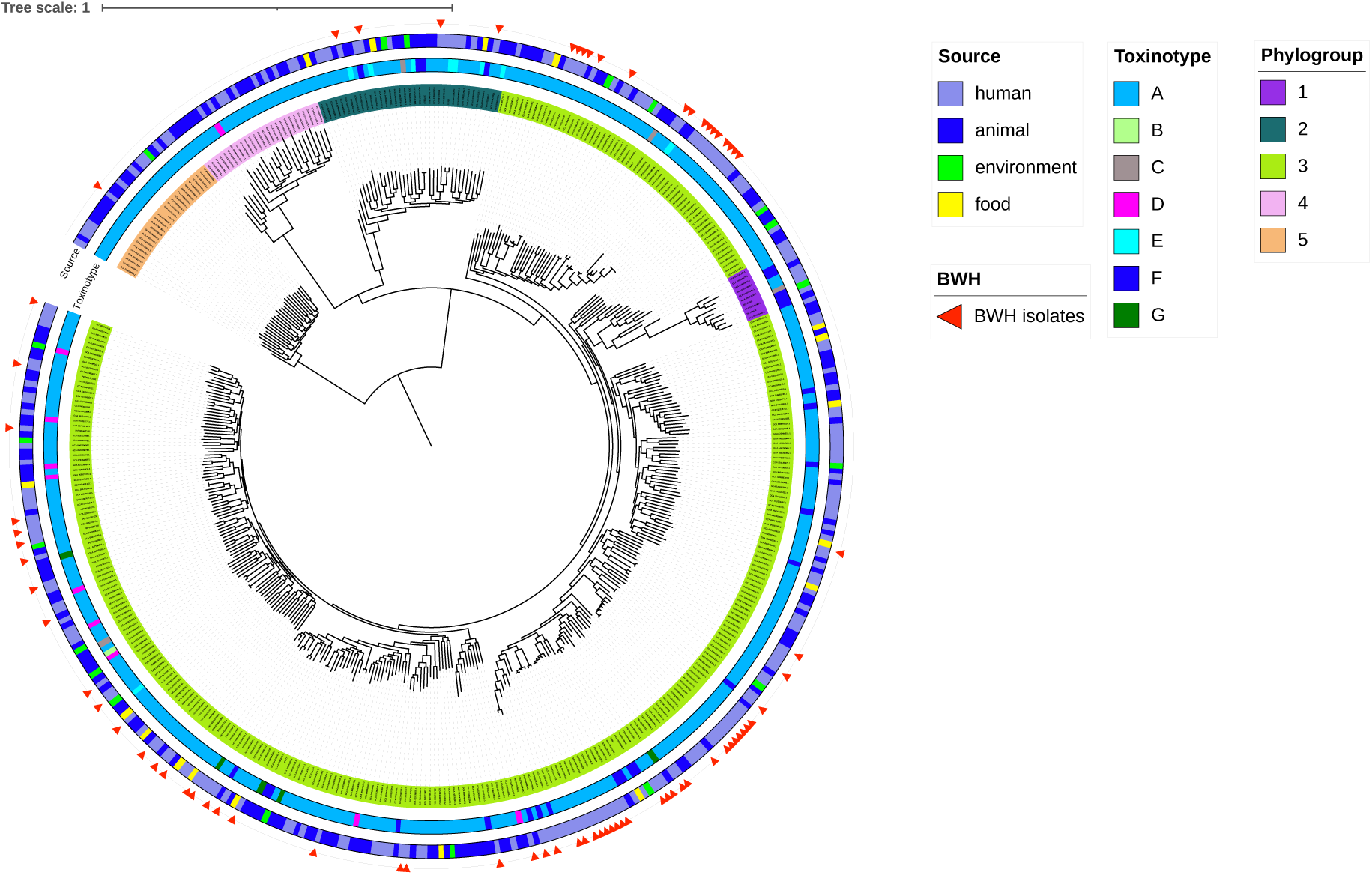
*Clostridium perfringens’* phylogenetic relationships. A maximum-likelihood (ML) phylogeny using IQ-TREE evaluates core genome SNPs among 401 representative genomes from NCBI Pathogen Detection. The key in the upper right-hand corner correlates with the outer to inner rings, noting BWH isolates (red triangles), source, toxinotype, and phylogroup for the innermost ring. Consistent with previous phylogenetic studies[9], phylogroup I is nested within phylogroup III.

**Table 2:** Distribution of the five major phylogroups according to the origins and toxinotypes of globally distributed *C. perfringens* sequences indexed at NCBI Pathogen Detection.

|  | Phylogroup |  |  |  |  |
| --- | --- | --- | --- | --- | --- |
|  | I | II | III | IV | V |
|  | 202 | 106 | 1912 | 45 | 56 |
| **Source** |  |  |  |  |  |
| **Human** | **111 (55%)** | 38 (35.8%) | 994 (52.0%) | 22 (48.9%) | 38 (67.9%) |
| **Animal** | 9 (4.5%) | **60 (56.6%)** | 737 (38.5%) | 23 (51.1%) | 15 (26.8%) |
| **Food** | **71 (35.1%)** | 5 (4.7%) | 95 (5%) | 0 (0%) | 0 (0%) |
| **Environment** | 11 (5.4%) | 3 (2.8%) | 86 (4.5%) | 0 (0%) | 3 (5.4%) |
| p value<sup>1</sup> | <0.0001 | 0.0006 | <0.0001 | 0.0338 | 0.0229 |
| **Toxinotype** |  |  |  |  |  |
| **A** | 32 (15.8%) | **56 (52.8%)** | **1549 (81%)** | **44 (97.8%)** | **56 (100%)** |
| **B** | 0 (0%) | 0 (0%) | 9 (0.5%) | 0 (0%) | 0 (0%) |
| **C** | 2 (1%) | 1 (0.9%) | **15 (0.8%)** | 0 (0%) | 0 (0%) |
| **D** | 0 (0%) | 0 (0%) | **55 (2.9%)** | 1 (2.2%) | 0 (0%) |
| **E** | 0 (0%) | **16 (15.1%)** | 2 (0.1%) | 0 (0%) | 0 (0%) |
| **F** | **168 (83.2%)** | **33 (31.1%)** | **211 (11%)** | 0 (0%) | 0 (0%) |
| **G** | 0 (0%) | 0 (0%) | **71 (3.7%)** | 0 (0%) | 0 (0%) |
| p value<sup>1</sup> | <0.0001 | <0.0001 | <0.0001 | 0.0086 | 0.0007 |

Genomes obtained from the NCBI Pathogen Detection *C. perfringens* resource. Data are presented as n (%).
<sup>1</sup> P-values were calculated using either the chi-square test or Fisher's exact test, as appropriate, to assess differences in toxinotype distribution across sources or phylogroups.

Notably, no clonal linkages were identified among the BWH patient isolates, indicating that foodborne or other nosocomial reservoirs of patient-to-patient transmission are unlikely. The majority of BWH isolates clustered within phylogroup III (n=69, 93%), 4 belonged to phylogroup II (5%), and one to phylogroup IV (Figure 1). All but one isolate fell within toxinotype A. Seventeen isolates (23%) clustered with known global SNP clusters, while the remainder were unique.

### BWH *C. perfringens* isolates display a distinct extraintestinal virulence profile

Among BWH patient isolates, analyses of 76 reported virulence genes showed a high prevalence (>90%) of hyaluronidase genes *nagHIJK*, sialidase genes *nanIJH*, and perfringolysin O (*pfoA*), mirroring the profiles observed among extraintestinal strains (Supplementary Table S4, Figure S3B). Moreover, comparative analysis of virulence gene distribution within the global collection indicated varying pathogenic potential across strains according to phylogroup, toxinotype, and source of isolation (Supplementary Table S4, Figure S3A).

Among the global phylogroups, I and V had the lowest virulence gene carriage, lacking the hyaluronidase genes (*nagHIJKL*) and sialidase *nanI* and *nanJ*, whereas phylogroup II was characterized by the absence of *cpb2*, *pfoA*, and iron uptake systems (Supplementary Table S4).

Virulence gene carriage also varied by source and associated toxinotypes. Food-derived isolates lacked key colonization and invasion genes (*nagHIJKL*, *nanI*, *nanJ*, *pfoA*), which were highly prevalent among extra-intestinal clinical isolates compared with intestinal isolates. Importantly, these differences remained significant after restricting the analysis to non-clonal strains, retaining only one representative per SNP cluster (Supplementary Table S4). Further pan-GWAS analyses correcting for population structure identified the sialidase NanI as the only virulence factor significantly associated with extra-intestinal isolates (β = 3.36; FDR = 0.049), consistent with moderate genomic inflation (*λ* = 1.2) (Supplementary Table S5).

### The hyluronidase *nagL* is associated with invasive extra-intestinal C. *perfringens* infections at BWH

Analyses of *C. perfringens* genes associated with severe invasive infections in immunocompromised patients identified an association with the *nagL* hyaluronidase (multivariate OR = 3.90, 95% CI = 1.14-16.24, p = 0.029). In contrast, *fbpA* toxin carriage showed no significant association (Table 3). Additional clinical covariates, strongly associated with invasiveness, included a high Charlson Comorbidity Index (OR = 1.23, 95% CI = 1.04-1.50, p = 0.013) and polymicrobial infection (OR = 0.04, 95% CI = 0.00-0.38, p = 0.0018) (Table 3).

**Table 3:**
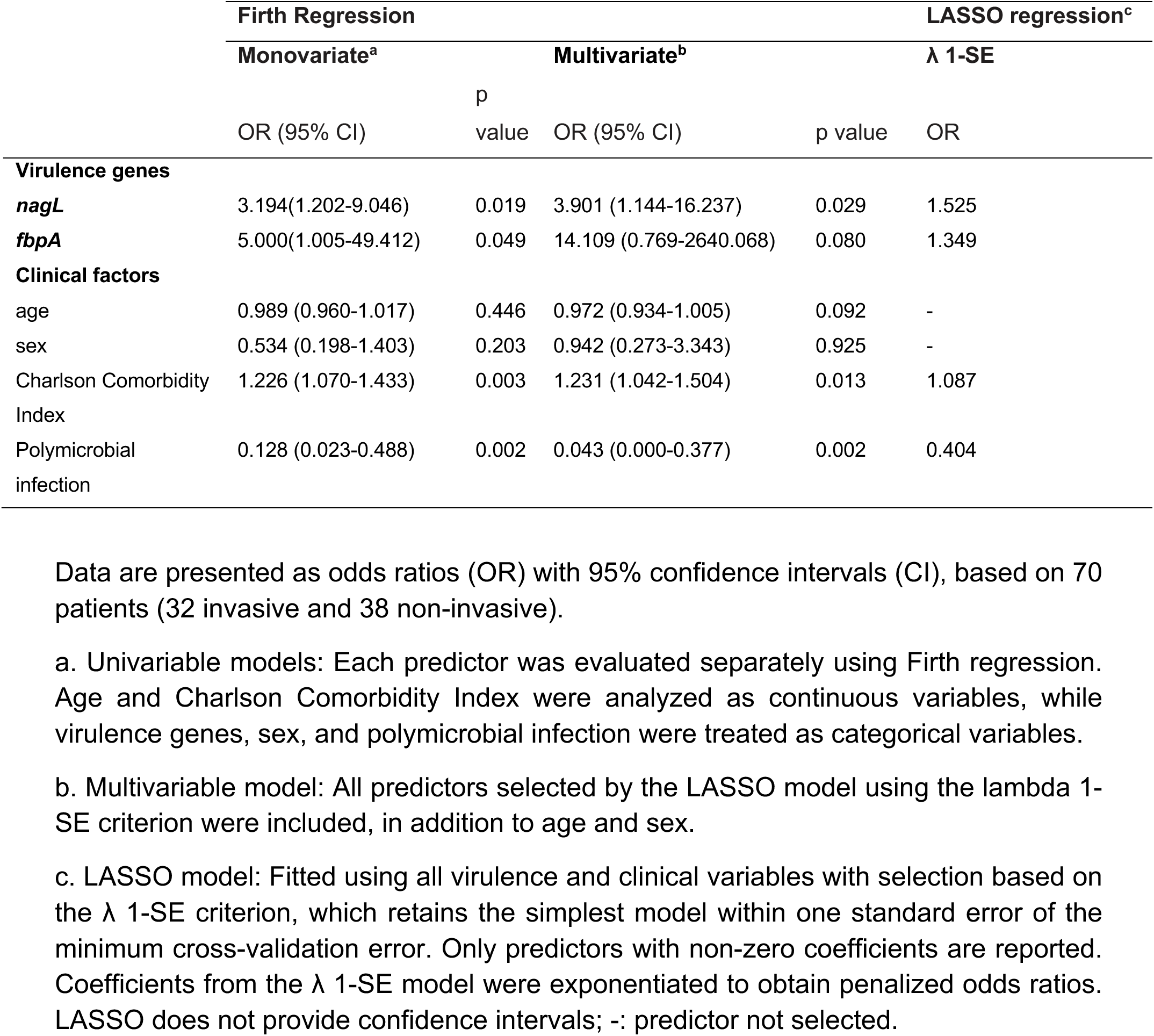
Association of *C. perfringens* virulence determinants with invasive infections among BWH patients.

### BWH isolates had high rates of carriage of tetracycline resistance genes but not against primary therapeutic agents, including macrolides or penicillins

Antimicrobial resistance (AMR) genes were detected in 74.4% (58/78) of the BWH genomes, with tetracycline resistance determinants *tetA* (74.4%) and *tetB* (56.4%) being the most prevalent, followed by the macrolide and lincosamide resistance gene *ermQ* (9%), lincosamide-resistance gene *lnuP* (5.1%), and aminoglycoside resistance genes *ant*(6)-Ib (2.6%) and *aac*(6’)-*Ie*/*aph*(2’’)-*Ia* (2.6%) (Supplementary Table S6). Among globally-deposited *C. perfringens*, except for *tetB*, no significant differences were observed in carriage of resistance genes between human extraintestinal and intestinal isolates (Supplementary Table S6).

## Discussion

We present genomic-epidemiologic analyses of *C. perfringens* cases in a large academic medical center, illustrating its emergence in highly morbid infections among cancer and other immunocompromised patients [3,5,6]. Nearly half of *C. perfringens* infections diagnosed within the hospital were invasive, particularly for sepsis, where mortality rates exceeded 40%, consistent with observations reported by other studies [3,6].

Our patient *C. perfringens* isolates were highly diverse and non-clonal, ruling-out nosocomial transmission and suggesting alternate reservoirs, such as the patients’ digestive tracts. Despite limited data on *C. perfringens* colonization, a recent Chinese surveillance study reported high carriage rates of *C. perfringens* in healthy individuals (45.8%) and ICU patients (12.8%) [25]. Taken together, hospitals may see constant reintroduction of diverse *C. perfringens* strains despite using best practices.

While the BWH patient strains belonged predominantly to phylogroup III and toxinotype A, they shared features with globally sequenced strains causing extraintestinal infections. Virulence gene analyses revealed higher prevalence of *pfoA* [26], the *nagHIJKL* hyaluronidases, *nanIJ* sialidase [27], and iron uptake systems [9] in extraintestinal isolates, consistent with their role in host invasion. Kiu et al. showed that hypovirulent intestinal strains in humans lack *pfoA*, in contrast to virulent lineages, corroborating the finding seen in infecting BWH isolates [28]. Assessments of larger cohorts may identify additional virulence genes and genomic factors associated with invasive infections in immunocompromised patients.

Our analyses revealed a strong association between *nagL* carriage and invasive infection, particularly in monomicrobial *C. perfringens* infections occurring in highly comorbid patients. NagL is part of a virulence network regulated by the VirS/VirR–RevR system and is co-regulated with major toxins (*plc*, *pfoA*, *colA*) and extracellular enzymes [2,29]. As shown by Andre et al. [30], cysteine availability modulates the expression of *nagL*, *pfoA*, and redox-related genes, linking sulfur metabolism to virulence regulation. Under cysteine-limited conditions in host tissues, this co-regulation likely promotes tissue invasion and damage in extraintestinal infections. Our discovery of its association with severe infections in co-morbid patients opens interesting questions regarding its expression during invasive infections, its co-expression with other toxins like *pfoA* and *cpa,* and its role in the context of impaired host immune responses.

Although antibiotic resistance has not significantly impacted treatment of *C. perfringens* infections, 74.4% of strains carried tetracycline resistance genes, and 14%, one or more macrolide and lincosamide resistance genes, consistent with previous reports [10]. While resistance gene acquisition in *C. perfringens* has been linked to antibiotic use in livestock, its presence in human clinical isolates raises concern for the emergence of resistant hospital strains. While *C. perfringens* remains largely susceptible to β-lactams, resistance to the lincosamide clindamycin is increasing (>20%), raising concern given its frequent use in penicillin-allergic patients [31].

Our study provides the first genomic-epidemiologic analysis of extraintestinal *C. perfringens* seen in patients who are immunocompromised or who harbor multiple co-morbidities, and who suffer high rates of mortality from these infections. Limitations of our study include its observational design and relatively small sample size, albeit of a relatively uncommon infection seen in hospital settings. Leveraging a broader, global repository of *C. perfringens* genomes, we identified multiple pathogen-associated factors, including hyaluronidase, sialidase, and *pfoA* carriage in extraintestinal isolates, and a new association of the NagL hyaluronidase with invasive infections, in addition to clinical factors that may modulate risks for these infections in highly vulnerable patient populations.

## Data availability

WGS data can be found under NCBI BioProject PRJNA420718.

## Acknowledgements/Funding

This work was supported by NIDDK grant P30 DK034854, the Massachusetts Life Sciences Center (Bry), the New York State Department of Health, and the Food and Drug Administration’s LFFM Grant (Cooperative Agreement number 1U19FD007089). We thank Dr. Michael Monuteaux for statistical support and Dr. Nicole Pecora and Jay Worley for their thoughtful insights and critical review of the manuscript. We thank the staff of the Advanced Genomic Technologies Cluster at the Wadsworth Center, where library preparation and sequencing were carried out. We thank the staff of the Clinical Microbiology Laboratory, Anaerobes section, BWH, where *C. perfringens* strains were isolated and identified.

