## Supplementary material for "Genomic and clinical determinants of extraintestinal *Clostridium perfringens* infections in immunocompromised patients": Online Supplement

### Table of contents

| | Page |
|---|---|
| **Table S1.** The STROBE checklist and additional STROME-ID items | 2-4 |
| **Table S2.** Distribution of the five major phylogroups according to the source of isolation and animal species | 5 |
| **Table S3.** Frequency distribution of C. perfringens sequence types (STs). | 6-7 |
| **Table S4.** Distribution of the virulence factors according to phylogroups, Toxinotypes, and sources | 8-10 |
| **Table S5.** Significant pan-GWAS associations with extra-intestinal *C. perfringens* isolates (Pyseer, FDR < 0.05) | 11-12 |
| **Table S6.** Distribution of the resistance genes according to phylogroups, Toxinotypes, and sources | 13 |
| **Figure S1.** GWAS power calculations by PowerBacGWAS for allele frequency and effect size combinations at the study sample size | 14 |
| **Figure S2.** Toxinotype distribution by phylogroup, source, and animal species | 15 |
| **Figure S3.** Virulence gene distribution by phylogroup, toxinotype, and source of isolation | 16 |

**Table S1**: The STROBE checklist and additional STROME-ID items

| | Item No | STROBE items | STROME-ID items | Line No |
|---|---|---|---|---|
| **Title and abstract** | 1 | (*a*) Indicate the study's design with a commonly used term in the title or the abstract | STROME-ID 1.1: The term molecular epidemiology should be applied to the study in the title or abstract and the keywords when molecular and epidemiological methods contribute substantially to the study | 21 |
| | | (*b*) Provide in the abstract an informative and balanced summary of what was done and what was found | | 21-34 |
| **Introduction** | | | | |
| Background/rationale | 2 | Explain the scientific background and rationale for the investigation being reported | STROME-ID 2.1: provide background information about the pathogen population and the distribution of pathogen strains within the host population at risk | 57-68 |
| Objectives | 3 | State specific objectives, including any prespecified hypotheses | STROME-ID 3.1: state the epidemiological objectives of using molecular typing | 77-82 |
| **Methods** | | | | |
| Study design | 4 | Present key elements of study design early in the paper | | 86-94 |
| | | Molecular terminology | STROME-ID 4.1: define or cite definitions for key molecular terms used within the study (eg,strain, isolate, and clone) | 119-124 |
| | | Molecular markers | STROME-ID 4.2: clearly define the molecular markers that were used with a standard nomenclature | 119-124 |
| | | Infectious disease case definition | STROME-ID 4.3: clearly state the infectious-disease case definitions | 93-94 |
| | | Laboratory methodology | STROME-ID 4.4: describe sample collection and laboratory methods, including any methods used to minimise and measure cross-contamination, and give the criteria used to interpret strain classification | 85-86; 100-102 |
| Setting | 5 | Describe the setting, locations, and relevant dates, including periods of recruitment, exposure, follow-up, and data collection | STROME-ID 5.1: clearly state the timeframe of the study; consider and appropriately reference the molecular clock of markers if known, and the natural history of the infection | 86-94 |
| Participants | 6 | (*a*) Give the eligibility criteria, and the sources and methods of case ascertainment and control selection. Give the rationale for the choice of cases and controls | STROME-ID 6.1: state the source of participants and clinical specimens, and clearly describe sampling frame and strategy | 86-97 |
| | | (*b*) For matched studies, give matching criteria and the number of controls per case | | Not applicable |
| Variables | 7 | Clearly define all outcomes, exposures, predictors, potential confounders, and effect modifiers. Give diagnostic criteria, if applicable | | 148-159 |
| Data sources/ measurement | 8* | For each variable of interest, give sources of data and details of methods of assessment (measurement). Describe comparability of assessment methods if there is more than one group | | Table 1 |
| | | | STROME-ID 8.1: describe any methods used to detect multiple-strain infections and measure their effect on the study findings | Not applicable |
| Bias | 9 | Describe any efforts to address potential sources of bias | STROME-ID 9.1: describe any efforts made to address discovery or ascertainment bias | 148-159 |
| Study size | 10 | Explain how the study size was arrived at | STROME-ID 10.1: describe any unique restrictions placed on the study sample size | 86-97 |

| | | | | |
|---|---|---|---|---|
| Quantitative variables | 11 | Explain how quantitative variables were handled in the analyses. If applicable, describe which groupings were chosen and why | | Not applicable |
| Statistical methods | 12 | (*a*) Describe all statistical methods, including those used to control for confounding | STROME-ID 12.1: state how the study took account of the non-independence of sample data, if appropriate<br>STROME-ID 12.2: state how the study dealt with missing data | 138-147; 156-157 |
| | | (*b*) Describe any methods used to examine subgroups and interactions | | Not applicable |
| | | (*c*) Explain how missing data were addressed | | 156-157 |
| | | (*d*) If applicable, explain how matching of cases and controls was addressed | | Not applicable |
| | | (*e*) Describe any sensitivity analyses | | Not applicable |
| **Results** | | | | |
| Participants | 13* | (a) Report numbers of individuals at each stage of study—eg numbers potentially eligible, examined for eligibility, confirmed eligible, included in the study, completing follow-up, and analysed | STROME-ID 13.1: Report numbers of participants and samples at each stage of the study, including the number of samples obtained, the number typed, and the number yielding data | 86-97; 165-167 |
| | | (b) Give reasons for non-participation at each stage | STROME-ID 13.2: if the study investigates groups of genetically indistinguishable pathogens (molecular clusters), state the sampling fraction, the distribution of cluster sizes, and the study population turnover, if known | Not applicable |
| | | (c) Consider use of a flow diagram | | Not applicable |
| Descriptive data | 14* | (a) Give characteristics of study participants (eg demographic, clinical, social) and information on exposures and potential confounders | STROME-ID 14.1: give information by strain type if appropriate, with use of standardised nomenclature | Table 1 |
| | | (b) Indicate number of participants with missing data for each variable of interest | | Table 1 |
| Outcome data | 15* | Report numbers in each exposure category, or summary measures of exposure | | Table 1 |
| Main results | 16 | (*a*) Give unadjusted estimates and, if applicable, confounder-adjusted estimates and their precision (eg, 95 confidence interval). Make clear which confounders were adjusted for and why they were included | STROME-ID 16.1: consider showing molecular relatedness of strain types by means of a dendrogram or phylogenetic tree | Table 3, Figure 1 |
| | | (*b*) Report category boundaries when continuous variables were categorized | | Not applicable |
| | | (*c*) If relevant, consider translating estimates of relative risk into absolute risk for a meaningful time period | | Not applicable |
| Other analyses | 17 | Report other analyses done—eg analyses of subgroups and interactions, and sensitivity analyses | | Not applicable |
| **Discussion** | | | | |
| Key results | 18 | Summarise key results with reference to study objectives | | 244-251; 255-257; 264-266 |
| Limitations | 19 | Discuss limitations of the study, taking into account sources of potential bias or imprecision. Discuss both direction and magnitude of any potential bias | STROME-ID 19.1: consider alternative explanations for findings when transmission chains are being investigated, and report the consistency between molecular and epidemiological evidence | 286-287 |

| | | | | |
|---|---|---|---|---|
| Interpretation | 20 | Give a cautious overall interpretation of results considering objectives, limitations, multiplicity of analyses, results from similar studies, and other relevant evidence | | 246-248<br>257-261<br>266-271 |
| Generalisability | 21 | Discuss the generalisability (external validity) of the study results | | 283-286 |
| **Other information** | | | | |
| Funding | 22 | Give the source of funding and the role of the funders for the present study and, if applicable, for the original study on which the present article is based | STROME-ID 23.1: report any ethical considerations with specific implications for infectious-disease molecular epidemiology | 354-356<br>87-89 |

**Table S2**: Distribution of the five major phylogroups according to the source of isolation and animal species

| | Phylogroup | | | | |
|---|---|---|---|---|---|
| | **I**<br>**202** | **II**<br>**106** | **III**<br>**1912** | **IV**<br>**45** | **V**<br>**56** |
| **Source of isolation** | | | | | |
| **ANIMAL** | 9 (4.5) | 60 (56.6) | 737 (38.5) | 23 (51.1) | 15 (26.8) |
| **Bovine** | 0 | 4 (3.8) | **98 (5.1)** | 0 | 4 (7.1) |
| **Goat** | 0 | 0 | 30 (1.6) | 0 | 0 |
| **Sheep** | 1 (0.5) | 1 (0.9) | 48 (2.6) | 1 (2.2) | 1 (1.8) |
| **Chicken** | 2 (1) | 4 (3.8) | **160 (8.3)** | 7 (15.6) | 2 (3.6) |
| **Bird** | 1 (0.5) | 0 (0) | 15 (0.8) | 3 (6.7) | 0 |
| **Dog** | 0 | **28 (26.4)** | 64 (3.3) | 1 (2.2) | 4 (7.1) |
| **Horse** | 0 | **11 (10.4)** | 5 (0.2) | 0 | 0 (0) |
| **Pet** | 0 | 4 (3.8) | 139 (7.3) | 4 (8.9) | 3 (5.4) |
| **Cat** | 0 | 3 (2.8) | 5 (0.3) | 2 (4.4) | 0 |
| **Pig** | 0 | 1 (0.9) | 74 (3.9) | 0 | 0 |
| **Other** | 0 | 4 (3.8) | 41 (2.1) | 2 (4.4) | 1 (1.8) |
| **NP** | 5 (2.5) | 0 | 58 (3) | 3 (6.7) | 0 |
| **HUMAN** | 111 (55) | 38 (35.8) | 994 (52.0) | 22 (48.9) | 38 (67.9) |
| **ENVIRONMENT** | 11 (5.4) | 3 (2.8) | 86 (4.5) | 0 | 3 (5.4) |
| **FOOD** | 71 (35.1) | 5 (4.7) | 95 (5) | 0 | 0 |
| p_value | 8.39e-63 | 0.00047 | 6.99e-21 | 0.0338 | 0.0229 |

Data are presented as n (%)
p-values compare frequencies across the sources of isolation within each phylogroup using $\chi^2$ or Fisher's exact tests, corrected by the Benjamini-Hochberg.

**Table S3**: Frequency distribution of *C. perfringens* sequence types (STs).

| Number of isolate per ST | Number of STs | ST list | Percentage in each ST (%) |
|---|---|---|---|
| **1229** | **-** | **Undetermined** | **52.95** |
| **36** | **1** | **248** | **1.55** |
| **28** | **1** | **72** | **1.21** |
| **25** | **1** | **21** | **1.08** |
| **24** | **1** | **41** | **1.03** |
| **23** | **1** | **73** | **0.99** |
| 22 | 1 | 353 | 0.95 |
| 21 | 1 | 77 | 0.9 |
| 20 | 2 | 139, 269 | 0.86 |
| 18 | 2 | 135, 147 | 0.78 |
| 17 | 4 | 154, 221, 36, 62 | 0.73 |
| 16 | 1 | 143 | 0.69 |
| 15 | 1 | 452 | 0.65 |
| 14 | 1 | 80 | 0.6 |
| 13 | 2 | 251, 308 | 0.56 |
| 12 | 1 | 335 | 0.52 |
| 11 | 4 | 102, 252, 274, 39 | 0.47 |
| 9 | 6 | 184, 250, 282, 29, 370, 78 | 0.39 |
| 8 | 3 | 142, 210, 33 | 0.34 |
| 7 | 7 | 132, 149, 176, 241, 285, 461, 5 | 0.3 |
| 6 | 6 | 144, 200, 312, 372, 43, 8 | 0.26 |
| 5 | 12 | 130, 32, 333, 367, 402, 408, 44, 479, 502, 538, 546, 566 | 0.22 |
| 4 | 8 | 207, 218, 222, 227, 316, 400, 42, 525 | 0.17 |
| 3 | 24 | 103, 151, 171, 172, 194, 240, 264, 278, 279, 298, 324, 340, 381, 389, 397, 410, 425, 431, 437, 45, 493, 521, 70, 71 | 0.13 |
| 2 | 66 | 104, 111, 112, 118, 119, 122, 13, 137, 150, 152, 162, 166, 175, 177, 179, 2, 216, 238, 239, 245, 246, 254, 255, 265, 268, 273, 287, 290, 294, 307, 31, 314, 315, 317, 322, 325, 326, 34, 344, 348, 354, 357, 369, 376, 388, 401, 421, 426, 430, 434, 450, 454, 457, 458, 466, 472, 485, 491, 500, 516, 532, 551, 558, 67, 83, 97 | 0.09 |
| **1** | **183** | 1, 100, 105, 106, 108, 109, 113, 114, 115, 124, 126, 127, 129, 134, 136, 138, 140, 141, 146, 153, 157, 158, 159, 160, 161, 163, 164, 165, 167, 169, 173, 178, 182, 183, 195, 206, 208, 209, 211, 215, 219, 226, 229, 231, 232, 235, 236, 237, 244, 247, 249, 25, 253, 256, 257, 258, 259, 260, 261, 262, 263, 267, 270, 271, 272, 275, 276, 280, 281, 283, 284, 286, 291, 292, 296, 299, 30, 309, 319, 323, 349, 35, 358, 383, 385, 396, 40, 404, 405, 409, 412, 413, 415, 417, 418, 420, 422, 423, 424, 428, 429, 432, 433, 435, 436, 439, 440, 441, 442, 443, 444, 445, 446, 447, 448, 449, 455, 456, 460, 462, 463, 464, 465, 467, 468, 469, 47, 470, 471, 473, 474, 475, 476, 477, 483, 484, 486, 487, 488, 489, 490, 492, 494, 498, 503, 506, 507, 508, 509, 512, 513, 514, 515, 518, 519, 52, 520, 523, 524, 526, 529, 53, 530, 533, 535, 55, 56, 563, 57, 59, 60, 61, 63, 68, 69, 74, 75, 76, 79, 82, 89, 93, 99 | 0.04 |

STs were grouped by the number of isolates in each ST. For every group, (i) the number of isolates per ST, (ii) the number of distinct STs with that count, (iii) the list of corresponding STs, and (iv) the percentage contribution of each ST to the full dataset (n = 2,321). “Undetermined” refers to isolates for which MLST typing could not be resolved from WGS data

**Table S4**: Distribution of virulence factors across globally available *C. perfringens* genomes (n = 2,321), stratified by phylogroup, toxinotype, and source of isolation.

| | Total | Phylogroup | | | | | | Toxinotype | | | | | | | |
|---|---|---|---|---|---|---|---|---|---|---|---|---|---|---|---|
| | 2231 | I 202 | II 106 | III 1912 | IV 45 | V 56 | p-value | A 1737 | F 412 | C 18 | B 9 | D 56 | G 71 | E 18 | p-value |
| **Typing toxins** | | | | | | | | | | | | | | | |
| *plc* | 2321 (100) | 202 (100) | 106 (100) | 1912 (100) | 45 (100) | 56 (100) | | 1737 (100) | 412 (100) | 18 (100) | 9 (100) | 56 (100) | 71 (100) | 18 (100) | |
| *cpe* | 441 (19) | **170 (84)** | **49 (46)** | 222 (12) | 0 | 0 | <0.0001 | 0 | **412 (100)** | 2 (11) | 0 | 9 (16) | 0 | 18 (100) | <0.0001 |
| *cpb* | 27 (1.2) | 2 (1) | 1 (0.9) | 24 (1.3) | 0 | 0 | | 0 | 0 | **18 (100)** | **9 (100)** | 0 | 0 | 0 | <0.0001 |
| *etx* | 65 (2.8) | 0 | 0 | **64 (3.3)** | 1 (2.2) | 0 | 0.0062 | 0 | 0 | 0 | **9 (100)** | **56 (100)** | 0 | 0 | <0.0001 |
| *netB* | 71 (3.1) | 0 | 0 | **71 (3.7)** | 0 | 0 | 0.0016 | 0 | 0 | 0 | 0 | 0 | **71 (100)** | 0 | <0.0001 |
| ***lap*** | 18 (0.8) | 0 | **16 (15)** | 2 (0.1) | 0 | 0 | <0.0001 | 0 | 0 | 0 | 0 | 0 | 0 | **18 (100)** | <0.0001 |
| *lbp* | 2 (0.1) | 0 | 0 | 2 (0.1) | 0 | 0 | | 0 | 0 | 0 | 0 | 0 | 0 | 2 (11) | 0.0002 |
| ***ibp_variant*** | 16 (0.7) | 0 | **16 (15)** | 0 | 0 | 0 | <0.0001 | 0 | 0 | 0 | 0 | 0 | 0 | **16 (89)** | <0.0001 |
| **Non-typing toxins and other virulence genes** | | | | | | | | | | | | | | | |
| ***becA*** | 35 (1.5) | 0 | **27 (26)** | 8 (0.4) | 0 | 0 | <0.0001 | 30 (1.7) | 3 (0.7) | 0 | 0 | 1 (1.8) | 0 | 1 (5.6) | |
| ***becB*** | 37 (1.6) | 0 | **27 (26)** | 10 (0.5) | 0 | 0 | <0.0001 | 32 (1.8) | 3 (0.7) | 0 | 0 | 1 (1.8) | 0 | 1 (5.6) | |
| ***edpA*** | 132 (5.7) | 1 (0.5) | 0 | **130 (6.8)** | 0 | 1 (1.8) | <0.0001 | **129 (7.4)** | 3 (0.7) | 0 | 0 | 0 | 0 | 0 | <0.0001 |
| *edpB* | 1 (0) | 0 | 0 | 1 (0.1) | 0 | 0 | | 1 (0.1) | 0 | 0 | 0 | 0 | 0 | 0 | |
| ***netE*** | 38 (1.6) | 0 | **26 (25)** | 12 (0.6) | 0 | 0 | <0.0001 | 1 (0.1) | **37 (9)** | 0 | 0 | 0 | 0 | 0 | <0.0001 |
| ***netF*** | 36 (1.6) | 0 | **25 (24)** | 11 (0.6) | 0 | 0 | <0.0001 | 1 (0.1) | **35 (8.5)** | 0 | 0 | 0 | 0 | 0 | <0.0001 |
| ***netG*** | 16 (0.7) | 0 | **16 (15)** | 0 | 0 | 0 | <0.0001 | 0 | **16 (3.9)** | 0 | 0 | 0 | 0 | 0 | <0.0001 |
| ***cnaA*** | 88 (3.8) | 2 (1) | 0 | 75 (3.9) | **11 (24)** | 0 | <0.0001 | 27 (1.6) | 0 | 2 (11) | 0 | 0 | **59 (83)** | 0 | <0.0001 |
| ***zmpA*** | 131 (5.6) | 0 | 0 | **129 (6.7)** | 2 (4.4) | 0 | <0.0001 | 61 (3.5) | 0 | 0 | 0 | 0 | **70 (99)** | 0 | <0.0001 |
| *zmpB* | 2243 (97) | 201 (96) | 106 (100) | 1852 (97) | 45 (100) | **39 (70)** | <0.0001 | 1660 (96) | 412 (100) | 18 (100) | 9 (100) | 55 (98) | 71 (100) | 18 (100) | <0.0001 |
| *Lam* | 11 (0.5) | 0 | 0 | 11 (0.6) | 0 | 0 | | 1 (0.1) | 0 | 1 (6) | **3 (33)** | **4 (7)** | 0 | **2 (11)** | <0.0001 |
| *cpb2atypical* | 1028 (44) | 3 (1.5) | 52 (49) | 929 (49) | 20 (44) | 24 (43) | <0.0001 | 781 (45) | 127 (31) | 3 (17) | 8 (89) | 29 (52) | 70 (99) | 10 (56) | <0.0001 |
| *cpb2consensus* | 184 (7.9) | 2 (1) | 0 | **182 (9.5)** | 0 | 0 | <0.0001 | 136 (7.8) | 41 (10) | 7 (39) | 0 | 0 | 0 | 0 | <0.0001 |
| *pfoA* | 1697 (73) | 0 | 102 (96) | 1539 (81) | **0** | 56 (100) | <0.0001 | 1314 (76) | **216 (52)** | 16 (89) | 9 (100) | 55 (98) | 70 (99) | 17 (94) | <0.0001 |
| *Delta* | 9 (0.4) | 0 | 1 (0.9) | 8 (0.4) | 0 | 0 | | 6 (0.3) | 2 (0.5) | 1 (5.6) | 0 | 0 | 0 | 0 | |
| *dlpA* | 5 (0.2) | 0 | 0 | 4 (0.2) | 1 (2.2) | 0 | | 4 (0.2) | 0 | 0 | 0 | 0 | 1 (1.4) | 0 | |
| *fbpA* | 2173 (94) | 202 (100) | 106 (100) | 1793 (94) | **34 (76)** | **38 (68)** | <0.0001 | 1616 (93) | 407 (99) | 14 (78) | 5 (56) | 43 (77) | 71 (100) | 17 (9) | <0.0001 |

| | Source | | | | | Human strains: origin | | | Human strains: origin (non clonal strains)* | | | BWH isolates | | |
|---|---|---|---|---|---|---|---|---|---|---|---|---|---|---|
| | human 1203 | animal 844 | food 171 | environment 103 | p-value | Intestinal 752 | Extra-intestinal 168 | p-value | Intestinal 368 | Extra-intestinal 154 | p-value | Non-invasive 38 | Invasive 32 | p-value |
| **Typing toxins** | | | | | | | | | | | | | | |
| *plc* | 1203 (100) | 844 (100) | 171 (100) | 103 (100) | | 752 (100) | 168 (100) | | 368 (100) | 154 (100) | | 38 (100) | 32 (100) | |
| *cpe* | 236 (20) | 90 (11) | **101 (59)** | 14 (14) | <0.0001 | **179 (24)** | 2 (1.2) | <0.0001 | **62 (17)** | 2 (1.3) | <0.0001 | 0 | 1 (3.1) | |
| *cpb* | 5 (0.4) | 20 (2.4) | 2 (1.2) | 0 | 0.0012 | 1 | 0 | | 1 (0.3) | 0 | | 0 | 0 | |
| *etx* | 5 (0.4) | **45 (5.3)** | 5 (2.9) | **10 (9.7)** | <0.0001 | 2 (0.3) | 0 | | 1 (0.3) | 0 | | 0 | 0 | |
| *netB* | 3 (0.2) | **65 (7.7)** | 3 (1.8) | 0 | <0.0001 | 3 (0.4) | 1 (0.6) | | 2 (0.5) | 1 (0.6) | | 0 | 0 | |
| ***lap*** | 6 (0.5) | **11 (1.3)** | 1 (0.6) | 0 | | 6 (0.8) | 0 | | 5 (1.4) | 0 | | 0 | 0 | |
| *lbp* | 0 | 2 (0.2) | 0 | 0 | | 0 | 0 | | 0 | 0 | | 0 | 0 | |
| ***ibp_variant*** | 6 (0.5) | 9 (1.1) | 1 (0.6) | 0 | | 6 (0.8) | 0 | | 5 (1.4) | 0 | | 0 | 0 | |
| **Non-typing toxins and other virulence genes** | | | | | | | | | | | | | | |
| ***becA*** | 25 (2.1) | 10 (1.2) | 0 | 0 | | 7 (1.9) | 2 (1.3) | | 16 (2.1) | 2 (1.2) | | 0 | 1 (3.1) | |
| ***becB*** | 25 (2.1) | 12 (1.4) | 0 | 0 | | 7 (1.9) | 2 (1.3) | | 16 (2.1) | 2 (1.2) | | 0 | 1 (3.1) | |
| ***edpA*** | **90 (7.5)** | 39 (4.6) | 3 (1.8) | 0 | <0.0001 | 76 (10) | 12 (7.1) | | 22 (6) | 11 (7.1) | | 3 (7.9) | 2 (6.2) | |
| *edpB* | 0 | 1 (0.1) | 0 | 0 | | 0 | 0 | | 0 | 0 | | 0 | 0 | |
| ***netE*** | 1 (0.1) | **38 (4.5)** | 0 | 0 | <0.0001 | 0 | 0 | | 0 | 0 | | 0 | 0 | |
| ***netF*** | 1 (0.1) | **36 (4.3)** | 0 | 0 | <0.0001 | 0 | 0 | | 0 | 0 | | 0 | 0 | |
| ***netG*** | 0 | **16 (1.9)** | 0 | 0 | <0.0001 | 0 | 0 | | 0 | 0 | | 0 | 0 | |
| ***cnaA*** | 15 (1.2) | **68 (8.1)** | 5 (2.9) | 0 | <0.0001 | 12 (1.6) | 1 (0.6) | | 9 (2.4) | 1 (0.6) | | 0 | 0 | |
| ***zmpA*** | 10 (0.8) | **108 (12.8)** | 13 (7.6) | 0 | <0.0001 | 7 (0.9) | 3 (1.8) | | 5 (1.4) | 2 (1.3) | | 0 | 2 (6.2) | |
| *zmpB* | 1173 (97) | 804 (95) | 170 (99) | 96 (93) | 0.0016 | 735 (98) | 165 (98) | | 353 (96) | 152 (99) | | 38 (100) | 30 (94) | |
| *Lam* | 1 (0.1) | **9 (1.1)** | 1 (0.6) | 0 | 0.0178 | 0 | 1 (0.6) | | 0 | 1 (0.6) | | 0 | 0 | |
| *cpb2atypical* | 534 (44) | 436 (52) | 36 (21) | 22 (21) | <0.0001 | 303 (40) | 83 (49) | | 164 (45) | 75 (49) | | 17 (45) | 12 (37) | |
| *cpb2consensus* | 84 (7) | 79 (9.4) | 17 (9.9) | 4 (3.9) | | 68 (9) | 6 (3.6) | | 29 (7.9) | 4 (2.6) | | 1 (2.6) | 0 | |
| *pfoA* | 814 (68) | 721 (85) | **85 (50)** | 77 (75) | <0.0001 | 461 (61) | **149 (89)** | <0.0001 | 231 (63) | **135 (88)** | | 37 (97) | 29 (91) | |
| *Delta* | 5 (0.4) | 4 (0.5) | 0 | 0 | | 3 (0.4) | 0 | | 3 (0.8) | 0 | | 0 | 0 | |
| *dlpA* | 0 | 4 (0.5) | 1 (0.6) | 0 | | 0 | 0 | | 0 | 0 | | 0 | 0 | |
| *fbpA* | 1156 (96) | 755 (89) | 167 (98) | 95 (92) | <0.0001 | 731 (97) | 154 (92) | 0.0045 | 353 (96) | 140 (91) | | 31 (82) | 31 (97) | |

| | | | | | | | | | | | | | | | | | | | | | | | | | | | | | |
|---|---|---|---|---|---|---|---|---|---|---|---|---|---|---|---|---|---|---|---|---|---|---|---|---|---|---|---|---|---|
| *fbpB* | 2321 (100) | 202 (100) | 106 (100) | 1912 (100) | 45 (100) | 56 (100) | | 1737 (100) | 412 (100) | 18 (100) | 9 (100) | 56 (100) | 71 (100) | 18 (100) | | 1203 (100) | 844 (100) | 171 (100) | 103 (100) | | 752 (100) | 168 (100) | | 368 (100) | 154 (100) | | 38 (100) | 32 (100) | |
| *ilpA* | 3 (0.1) | 0 | 0 | 1 (0.1) | 2 (4.4) | 0 | | 3 (0.2) | 0 | 0 | 0 | 0 | 0 | 0 | | 0 | 3 (0.4) | 0 | 0 | | 0 | 0 | | 0 | 0 | | 0 | 0 | |
| *ilpB* | 3 (0.1) | 0 | 0 | 1 (0.1) | 2 (4.4) | 0 | | 3 (0.2) | 0 | 0 | 0 | 0 | 0 | 0 | | 0 | 3 (0.4) | 0 | 0 | | 0 | 0 | | 0 | 0 | | 0 | 0 | |
| *ldpA* | 3 (0.1) | 0 | 0 | 0 | 3 (6.7) | 0 | | 2 (0.1) | 0 | 0 | 0 | 1 (1.8) | 0 | 0 | | 1 (0.1) | 2 (0.2) | 0 | 0 | | 0 | 0 | | 0 | 0 | | 0 | 0 | |
| *ldpB* | 8 (0.3) | 0 | 0 | 8 (0.4) | 0 | 0 | | 5 (0.3) | 0 | 0 | 0 | 0 | 3 (4.2) | 0 | 0.0207 | 1 (0.1) | 6 (0.7) | 1 (0.6) | 0 | | 1 (0.1) | 0 | | 1 (0.3) | 0 | | 0 | 0 | |
| *ldpC* | 27 (1.2) | 0 | 0 | 27 (1.4) | 0 | 0 | | 25 (1.4) | 0 | 0 | 0 | 1 (1.8) | 1 (1.4) | 0 | | 0 | 25 (3) | 2 (1.2) | 0 | <0.0001 | 0 | 0 | | 0 | 0 | | 0 | 0 | |
| *nagH* | 2263 (98) | 202 (100) | 106 (100) | 1910 (99) | 45 (100) | **0** | <0.0001 | 1679 (97) | 412 (100) | 18 (100) | 9 (100) | 56 (100) | 71 (100) | 18 (100) | 0.0006 | 1165 (97) | 827 (98) | 171 (100) | 100 (9) | | 722 (96) | 165 (98) | | 351 (95) | 151 (98) | | 37 (97) | 32 (100) | |
| *nagI* | 1977 (85) | **1 (0.5)** | 106 (100) | 1835 (96) | 35 (78) | **0 (0)** | <0.0001 | 1573 (91) | **235 (57)** | 15 (83) | 9 (100) | 56 (100) | 71 (100) | 18 (100) | <0.0001 | 998 (83) | 798 (95) | **93 (54)** | 88 (85) | <0.0001 | 595 (79) | **159 (95)** | <0.0001 | 300 (81) | **145 (94)** | 0.0013 | 37 (97) | 32 (100) | |
| *nagJ* | 2082 (90) | **45 (22)** | 106 (100) | 1886 (99) | 45 (100) | **0 (0)** | <0.0001 | 1634 (94) | **276 (67)** | 18 (100) | 9 (100) | 56 (100) | 71 (100) | 18 (100) | <0.0001 | 1082 (90) | 806 (95) | **112 (65)** | 82 (80) | <0.0001 | 663 (88) | **162 (96)** | 0.0065 | 327 (89) | **148 (96)** | 0.0376 | 37 (97) | 32 (100) | |
| *nagK* | 1877 (81) | **0 (0)** | 106 (100) | 1741 (91) | 30 (67) | **0 (0)** | <0.0001 | 1487 (86) | **232 (56)** | 9 (50) | 7 (78) | 53 (95) | 71 (100) | 18 (100) | <0.0001 | 980 (81) | 718 (85) | **98 (57)** | 81 (79) | <0.0001 | 607 (81) | **150 (89)** | 0.0313 | 299 (81) | **139 (90)** | 0.0404 | 36 (95) | 29 (91) | |
| *nagL* | 1332 (57) | **20 (9.9)** | 105 (99) | 1162 (61) | 45 (100) | **0 (0)** | <0.0001 | 1001 (58) | 238 (58) | 8 (44) | 9 (100) | 21 (37) | 39 (55) | 16 (89) | 0.0004 | 717 (60) | 498 (59) | 72 (42) | 45 (44) | <0.0001 | 453 (60) | 95 (56) | | 239 (65) | 87 (56) | | 18 (47) | **24 (75)** | **0.035** |
| *nanH* | 2319 (99) | 202 (100) | 106 (100) | 1910 (99) | 45 (100) | 56 (100) | | 1735 (99) | 412 (100) | 18 (100) | 9 (100) | 56 (100) | 71 (100) | 18 (100) | | 1202 (99) | 844 (100) | 171 (100) | 102 (99) | | 752 (100) | 167 (99) | | 368 (100) | 153 (99) | | 38 (100) | 32 (100) | |
| *nanI* | 1979 (85) | **7 (3.5)** | 106 (100) | 1781 (93) | 45 (100) | **40 (71)** | <0.0001 | 1583 (91) | **227 (55)** | 15 (8) | 9 (100) | 56 (100) | 71 (100) | 18 (100) | <0.0001 | 1029 (85) | 782 (93) | **92 (54)** | 76 (74) | <0.0001 | 614 (82) | **166 (99)** | <0.0001 | 310 (84) | **152 (99)** | <0.0001 | 38 (100) | 32 (100) | |
| *nanJ* | 2028 (87) | **44 (22)** | 106 (100) | 1833 (96) | 45 (100) | **0 (0)** | <0.0001 | 1581 (91) | **276 (67)** | 18 (100) | 9 (100) | 55 (98) | 71 (100) | 18 (100) | <0.0001 | 1051 (87) | 787 (93) | **112 (65)** | 78 (76) | <0.0001 | 637 (85) | **160 (95)** | 0.0015 | 313 (85) | **146 (95)** | 0.0008 | 37 (97) | 32 (100) | |
| *Perfrin* | 36 (1.6) | 0 | 1 (0.9) | 35 (1.8) | 0 | 0 | | 7 (0.4) | 0 | 0 | 0 | 0 | 27 (38) | 2 (11) | <0.0001 | 0 | **36 (4.3)** | 0 | 0 | <0.0001 | 0 | 0 | | 0 | 0 | | 0 | 0 | |
| *TpeL* | 42 (1.8) | 0 | 1 (0.9) | 41 (2.1) | 0 | 0 | | 2 (0.1) | 0 | 13 (72) | 9 (100) | 0 | 18 (25) | 0 | <0.0001 | 3 (0.2) | **37 (4.4)** | 2 (1.2) | 0 | <0.0001 | 0 | 0 | | 0 | 0 | | 0 | 0 | |
| *NCTC8081_02938* | 2 (0.1) | 2 (1) | 0 | 0 | 0 | 0 | | 0 | 0 | 2 (11) | 0 | 0 | 0 | 0 | | 2 (0.2) | 0 | 0 | 0 | | 0 | 0 | | 0 | 0 | | 0 | 0 | |
| *colA* | 2321 (100) | 202 (100) | 106 (100) | 1912 (100) | 45 (100) | 56 (100) | | 1737 (100) | 412 (100) | 18 (100) | 9 (100) | 56 (100) | 71 (100) | 18 (100) | | 1203 (100) | 844 (100) | 171 (100) | 103 (100) | | 752 (100) | 168 (100) | | 368 (100) | 154 (100) | | 38 (100) | 32 (100) | |
| *cloSI* | 2316 (99) | 202 (100) | 106 (100) | 1907 (99) | 45 (100) | 56 (100) | | 1734 (99) | 412 (100) | 17 (94) | 8 (89) | 56 (100) | 71 (100) | 18 (100) | 0.0083 | 1201 (99.8) | 842 (99.8) | 171 (100) | 102 (99) | | 752 (100) | 168 (100) | | 368 (100) | 154 (100) | | 38 (100) | 32 (100) | |
| **(Putative) Iron uptake systems** | | | | | | | | | | | | | | | | | | | | | | | | | | | | | |
| CPE_RS01460 | 2319 (99) | 202 (100) | 106 (100) | 1910 (99) | 45 (100) | 56 (100) | | 1735 (99) | 412 (100) | 18 (100) | 9 (100) | 56 (100) | 71 (100) | 18 (100) | | 1201 (99) | 844 (100) | 171 (100) | 103 (100) | | 752 (100) | 168 (100) | | 368 (100) | 154 (100) | | 38 (100) | 32 (100) | |
| CPE_RS01465 | 2071 (89) | 202 (100) | 104 (98) | **1666 (87)** | 45 (100) | 54 (96) | <0.0001 | **1558 (90)** | **341 (83)** | 18 (100) | 9 (100) | 56 (100) | 71 (100) | 18 (100) | <0.0001 | 1051 (87) | 778 (92) | 141 (82) | 101 (9) | <0.0001 | 627 (83) | **162 (98)** | <0.0001 | 315 (86) | **150 (97)** | 0.0006 | 38 (100) | 32 (100) | |
| CPE_RS01470 | 2071 (89) | 202 (100) | 104 (98) | **1666 (87)** | 45 (100) | 54 (96) | <0.0001 | **1558 (90)** | **341 (83)** | 18 (100) | 9 (100) | 56 (100) | 71 (100) | 18 (100) | <0.0001 | 1051 (87) | 778 (92) | 141 (82) | 101 (98) | <0.0001 | 627 (83) | **162 (98)** | <0.0001 | 315 (86) | **150 (97)** | 0.0006 | 38 (100) | 32 (100) | |
| CPE_RS01475 | 2071 (89) | 202 (100) | 104 (98) | **1666 (87)** | 45 (100) | 54 (96) | <0.0001 | **1558 (90)** | **341 (83)** | 18 (100) | 9 (100) | 56 (100) | 71 (100) | 18 (100) | <0.0001 | 1051 (87) | 778 (92) | 141 (82) | 101 (98) | <0.0001 | 627 (83) | **162 (98)** | <0.0001 | 315 (86) | **150 (97)** | 0.0006 | 38 (100) | 32 (100) | |
| CPE_RS01480 | 2071 (89) | 202 (100) | 104 (98) | **1666 (87)** | 45 (100) | 54 (96) | <0.0001 | **1558 (90)** | **341 (83)** | 18 (100) | 9 (100) | 56 (100) | 71 (100) | 18 (100) | <0.0001 | 1051 (87) | 778 (92) | 141 (82) | 101 (98) | <0.0001 | 627 (83) | **162 (98)** | <0.0001 | 315 (86) | **150 (97)** | 0.0006 | 38 (100) | 32 (100) | |
| CPE_RS01485 | 2071 (89) | 202 (100) | 104 (98) | **1666 (87)** | 45 (100) | 54 (9) | <0.0001 | **1558 (90)** | **341 (83)** | 18 (100) | 9 (100) | 56 (100) | 71 (100) | 18 (100) | <0.0001 | 1051 (87) | 778 (92) | 141 (82) | 101 (98) | <0.0001 | 627 (83) | **162 (98)** | <0.0001 | 315 (86) | **150 (97)** | 0.0006 | 38 (100) | 32 (100) | |
| CPE_RS01490 | 2071 (89) | 202 (100) | 104 (98) | **1666 (87)** | 45 (100) | 54 (96) | <0.0001 | **1558 (90)** | **341 (83)** | 18 (100) | 9 (100) | 56 (100) | 71 (100) | 18 (100) | <0.0001 | 1051 (87) | 778 (92) | 141 (82) | 101 (98) | <0.0001 | 627 (83) | **162 (98)** | <0.0001 | 315 (86) | **150 (97)** | 0.0006 | 38 (100) | 32 (100) | |
| CPE_RS01495 | 2319 (99) | 202 (100) | 106 (100) | 1910 (99) | 45 (100) | 56 (100) | | 1735 (99) | 412 (100) | 18 (100) | 9 (100) | 56 (100) | 71 (100) | 18 (100) | | 1201 (99) | 844 (100) | 171 (100) | 103 (100) | | 752 (100) | 168 (100) | | 368 (100) | 154 (100) | | 38 (100) | 32 (100) | |
| CPE_RS01500 | 2319 (99) | 202 (100) | 106 (100) | 1910 (99) | 45 (100) | 56 (100) | | 1735 (99) | 412 (100) | 18 (100) | 9 (100) | 56 (100) | 71 (100) | 18 (100) | | 1201 (99) | 844 (100) | 171 (100) | 103 (100) | | 752 (100) | 168 (100) | | 368 (100) | 154 (100) | | 38 (100) | 32 (100) | |
| CPE_RS02585 | 1741 (75) | **0** | 55 (52) | 1586 (93) | 45 (100) | 55 (98) | <0.0001 | 1391 (80) | 210 (51) | 15 (83) | 9 (100) | 56 (100) | 56 (79) | 4 (22) | <0.0001 | 874 (73) | 710 (84) | 89 (52) | 68 (66) | <0.0001 | 550 (73) | **145 (86)** | 0.0015 | 264 (72) | **132 (86)** | 0.0031 | 35 (92) | 28 (87) | |
| CPE_RS02590 | 1742 (75) | **0** | 55 (52) | 1587 (83) | 45 (100) | 55 (98) | <0.0001 | 1392 (80) | 210 (51) | 15 (83) | 9 (100) | 56 (100) | 56 (79) | 4 (22) | <0.0001 | 875 (73) | 710 (84) | 89 (52) | 68 (66) | <0.0001 | 550 (73) | **146 (89)** | 0.0009 | 264 (72) | **133 (86)** | 0.0019 | 35 (92) | 28 (87) | |
| CPE_RS02595 | 1741 (75) | **0** | 55 (52) | 1585 (83) | 45 (100) | 55 (98) | <0.0001 | 1390 (80) | 210 (51) | 15 (83) | 9 (100) | 56 (100) | 56 (79) | 4 (22) | <0.0001 | 874 (73) | 709 (84) | 89 (52) | 68 (66) | <0.0001 | 550 (73) | **145 (86)** | 0.0015 | 264 (72) | **132 (86)** | 0.00312 | 35 (92) | 28 (87) | |

| | | | | | | | | | | | | | | | | | | | | | | | | | | | | | |
|---|---|---|---|---|---|---|---|---|---|---|---|---|---|---|---|---|---|---|---|---|---|---|---|---|---|---|---|---|---|
| CPE_RS04350 | 1481 (64) | 175 (87) | 97 (92) | 1200 (63) | **9 (20)** | **0** | <0.0001 | 1018 (59) | 327 (79) | 16 (89) | 7 (79) | 31 (55) | 65 (9) | 17 (94) | <0.0001 | 807 (67) | 512 (612) | 116 (68) | 46 (45) | <0.0001 | 496 (66) | 115 (68) | | 243 (66) | 104 (67) | | 26 (68) | 21 (66) | |
| CPE_RS04355 | 1481 (64) | 175 (87) | 97 (9) | 1200 (63) | **9 (20)** | **0** | <0.0001 | 1018 (59) | 327 (79) | 16 (89) | 7 (78) | 31 (55) | 65 (91) | 17 (94) | <0.0001 | 807 (67) | 512 (60) | 116 (68) | 46 (45) | <0.0001 | 496 (66) | 115 (68) | | 243 (66) | 104 (67) | | 26 (68) | 21 (66) | |
| CPE_RS04360 | 1481 (64) | 175 (87) | 97 (91) | 1200 (63) | **9 (20)** | **0** | <0.0001 | 1018 (59) | 327 (79) | 16 (89) | 7 (78) | 31 (55) | 65 (91) | 17 (94) | <0.0001 | 807 (67) | 512 (60) | 116 (68) | 46 (45) | <0.0001 | 496 (66) | 115 (68) | | 243 (66) | 104 (67) | | 26 (68) | 21 (66) | |
| CPE_RS04365 | 1482 (64) | 175 (87) | 97 (91) | 1201 (63) | **9 (20)** | **0** | <0.0001 | 1019 (59) | 327 (79) | 16 (89) | 7 (78) | 31 (55) | 65 (91) | 17 (94) | <0.0001 | 808 (67) | 512 (61) | 116 (68) | 46 (45) | <0.0001 | 496 (66) | 115 (68) | | 243 (66) | 104 (67) | | 26 (68) | 21 (66) | |
| CPE_RS04435 | 2286 (98) | 170 (84) | 104 (98) | 1911 (99) | 45 (100) | 56 (100) | <0.0001 | 1734 (9) | 380 (92) | 18 (100) | 9 (100) | 56 (100) | 71 (100) | 18 (100) | <0.0001 | 1187 (99) | 840 (99) | 156 (91) | 103 (100) | <0.0001 | 737 (98) | 168 (100) | | 363 (99) | 154 (100) | | 38 (100) | 32 (100) | |
| CPE_RS04440 | 2286 (98) | 170 (84) | 104 (98) | 1911 (99) | 45 (100) | 56 (100) | <0.0001 | 1734 (9) | 380 (92) | 18 (100) | 9 (100) | 56 (100) | 71 (100) | 18 (100) | <0.0001 | 1187 (99) | 840 (99) | 156 (91) | 103 (100) | <0.0001 | 737 (98) | 168 (100) | | 363 (99) | 154 (100) | | 38 (100) | 32 (100) | |
| CPE_RS04445 | 2286 (98) | 170 (84) | 104 (98) | 1911 (99) | 45 (100) | 56 (100) | <0.0001 | 1734 (99) | 380 (92) | 18 (100) | 9 (100) | 56 (100) | 71 (100) | 18 (100) | <0.0001 | 1187 (99) | 840 (99) | 156 (91) | 103 (100) | <0.0001 | 737 (98) | 168 (100) | | 363 (98.6%) | 154 (100) | | 38 (100) | 32 (100) | |
| CPE_RS05550 | 2318 (99) | 202 (100) | 106 (100) | 1909 (98) | 45 (100) | 56 (100) | | 1734 (99) | 412 (100) | 18 (100) | 9 (100) | 56 (100) | 71 (100) | 18 (100) | | 1200 (99) | 844 (100) | 171 (100) | 103 (100) | | 749 (99) | 168 (100) | | 368 (100) | 154 (100) | | 38 (100) | 32 (100) | |
| CPE_RS05555 | 2318 (99) | 202 (100) | 106 (100) | 1909 (99) | 45 (100) | 56 (100) | | 1734 (99) | 412 (100) | 18 (100) | 9 (100) | 56 (100) | 71 (100) | 18 (100) | | 1200 (99) | 844 (100) | 171 (100) | 103 (100) | | 749 (99) | 168 (100) | | 368 (100) | 154 (100) | | 38 (100) | 32 (100) | |
| CPE_RS05560 | 2318 (99) | 202 (100) | 106 (100) | 1909 (99) | 45 (100) | 56 (100) | | 1734 (99) | 412 (100) | 18 (100) | 9 (100) | 56 (100) | 71 (100) | 18 (100) | | 1200 (99) | 844 (100) | 171 (100) | 103 (100) | | 749 (99) | 168 (100) | | 368 (100) | 154 (100) | | 38 (100) | 32 (100) | |
| CPE_RS06390 | 1904 (82) | **3 (1.5)** | 106 (100) | 1695 (88) | 45 (100) | 55 (98) | <0.0001 | 1556 (90) | 178 (43) | 16 (89) | 9 (100) | 56 (100) | 71 (100) | 18 (100) | <0.0001 | 976 (81) | 773 (97) | 72 (4) | 83 (81) | <0.0001 | 578 (77) | **161 (96)** | <0.0001 | 293 (80) | **147 (95)** | <0.0001 | 35 (92) | 28 (87) | |
| CPE_RS06395 | 1904 (82) | **3 (1.5)** | 106 (100) | 1695 (88) | 45 (100) | 55 (98) | <0.0001 | 1556 (90) | 178 (43) | 16 (89) | 9 (100) | 56 (100) | 71 (100) | 18 (100) | <0.0001 | 976 (81) | 773 (97) | 72 (42) | 83 (81) | <0.0001 | 578 (77) | **161 (96)** | <0.0001 | 293 (80) | **147 (95)** | <0.0001 | 35 (92) | 28 (87) | |
| CPE_RS06400 | 1905 (82) | **3 (1.5)** | 106 (100) | 1696 (88) | 45 (100) | 55 (98) | <0.0001 | 1557 (90) | 178 (43) | 16 (89) | 9 (100) | 56 (100) | 71 (100) | 18 (100) | <0.0001 | 976 (81) | 773 (92) | 73 (43) | 83 (81) | <0.0001 | 578 (77) | **161 (96)** | <0.0001 | 293 (80) | **147 (95)** | <0.0001 | 38 (100) | 31 (97) | |
| CPE_RS08555 | 2321 (100) | 202 (100) | 106 (100) | 1912 (100) | 45 (100) | 56 (100) | | 1737 (100) | 412 (100) | 18 (100) | 9 (100) | 56 (100) | 71 (100) | 18 (100) | | 1203 (100) | 844 (100) | 171 (100) | 103 (100) | | 752 (100) | 168 (100) | | 368 (100) | 154 (100) | | 38 (100) | 32 (100) | |
| CPE_RS08560 | 2321 (100) | 202 (100) | 106 (100) | 1912 (100) | 45 (100) | 56 (100) | | 1737 (100) | 412 (100) | 18 (100) | 9 (100) | 56 (100) | 71 (100) | 18 (100) | | 1203 (100) | 844 (100) | 171 (100) | 103 (100) | | 752 (100) | 168 (100) | | 368 (100) | 154 (100) | | 38 (100) | 32 (100) | |
| CPF_1009 | 2315 (99) | 201 (99) | 106 (100) | 1907 (99) | 45 (100) | 56 (100) | | 1733 (99) | 411 (99) | 17 (94) | 9 (100) | 56 (100) | 71 (100) | 18 (100) | | 1203 (100) | 840 (99) | 170 (99) | 102 (99) | | 752 (100) | 168 (100) | | 368 (100) | 154 (100) | | 38 (100) | 32 (100) | |
| CPF_1010 | 2315 (99) | 201 (99) | 106 (100) | 1906 (99) | 45 (100) | 56 (100) | | 1732 (99) | 411 (99) | 17 (94) | 9 (100) | 56 (100) | 71 (100) | 18 (100) | | 1203 (100) | 839 (99) | 170 (99) | 102 (99) | | 752 (100) | 168 (100) | | 368 (100) | 154 (100) | | 38 (100) | 32 (100) | |
| CPF_1125 | 2277 (98) | **164 (81)** | 104 (98) | 1908 (99) | 45 (100) | 56 (100) | <0.0001 | 1722 (99) | 385 (93) | 17 (94) | 9 (100) | 55 (98) | 71 (100) | 18 (100) | <0.0001 | 1180 (98) | 838 (99) | 157 (92) | 102 (99) | <0.0001 | 752 (100) | 168 (100) | | 364 (99) | 154 (100) | | 38 (100) | 32 (100) | |
| CPF_1126 | 2267 (97) | **164 (81)** | 104 (98) | 1899 (99) | 45 (100) | 55 (98) | <0.0001 | 1712 (98) | 385 (93) | 17 (94) | 9 (100) | 55 (98) | 71 (100) | 18 (100) | <0.0001 | 1179 (98) | 830 (98) | 156 (91) | 102 (99) | <0.0001 | 732 (97) | 167 (99) | | 364 (99) | 153 (99) | | 38 (100) | 32 (100) | |
| CPF_1127 | 2267 (97) | **164 (81)** | 104 (98) | 1899 (99) | 45 (100) | 55 (98) | <0.0001 | 1712 (98) | 385 (93) | 17 (94) | 9 (100) | 55 (98) | 71 (100) | 18 (100) | <0.0001 | 1179 (98) | 830 (98) | 156 (91) | 102 (99) | <0.0001 | 732 (97) | 167 (99) | | 364 (99) | 153 (99) | | 38 (100) | 32 (100) | |
| **Mean count$^{\lambda}$** | **43.0 (5.4)** | **33.9 (2.4)** | **47.4 (3.2)** | **44.0 (4.7)** | **42.9 (2.2)** | **36.2 (2.0)** | **<0.0001** | **43.2 (4.7)** | **40.3 (7.2)** | **46.4 (3.4)** | **49.6 (1.7)** | **46.0 (2.3)** | **50.1 (1.8)** | **48.1 (1.5)** | **<0.0001** | **42.8 (5.3)** | **44.5 (4.9)** | **38.9 (6.2)** | **41.4 (5.1)** | **<0.0001** | **42.3 (5.6)** | **44.8 (3.5)** | **<0.0001** | **42.0 (5.0)** | **44.2 (3.4)** | **<0.0001** | **45.1 (2.8)** | **45.0 (2.9)** | 0.99 |

Data shown as n ().
p-values compare frequencies across categories using $\chi^2$ or Fisher's exact tests, corrected by the Benjamini-Hochberg. Coloured cells highlight significant association of virulence factors with specific phylogroups, toxinotypes, or sources. Bold values indicate the most notable significant within-group differences
*: Only non-clonal human isolates were included, with one representative retained per SNP cluster to exclude clonal isolates.
$^{\lambda}$: Mean numbers of virulence genes per isolate were compared across groups using the Kruskal-Wallis test or Wilcoxon rank-sum, as appropriate

**Table S5.** Significant pan-GWAS associations with extra-intestinal *C. perfringens* isolates (Pyseer, FDR < 0.05)

| Variant | Annotation | af | lrt-pvalue | beta | beta-SE | intercept | FDR | PC1 | PC2 | PC3 | PC4 | PC5 | PC6 | PC7 | PC8 | PC9 | PC10 | PC11 | PC12 | PC13 | PC14 | PC15 | PC16 | PC17 | PC18 | PC19 | PC20 |
|---|---|---|---|---|---|---|---|---|---|---|---|---|---|---|---|---|---|---|---|---|---|---|---|---|---|---|---|
| **group_2117** | hypothetical proteins | 0.94 | 1.2e-10 | -2.43 | 0.40 | 0.24 | **<0.01** | -1.2 | 1.4 | 1.3 | 1.8 | 0.8 | 1.1 | -1.2 | -0.1 | -1.7 | -0.9 | -0.8 | -0.6 | 0.1 | -0.4 | -0.9 | 0.5 | -0.7 | 0.5 | -0.6 | 0.7 |
| **RubR2.1,RubR2.2,RubR2.3** | Rubrerythrin | 0.91 | 0.0005 | -1.3 | 0.39 | -0.77 | **0.017** | -1.2 | 1.5 | 0.9 | 1.5 | 0.7 | 1.0 | -0.7 | 0.4 | -0.7 | -0.6 | -0.9 | -0.3 | -0.2 | -0.3 | -1.11 | 0.4 | -0.6 | 0.6 | -0.2 | 1.0 |
| **rubR2_2** | Rubrerythrin | 0.88 | 0.0005 | -1.41 | 0.42 | -0.719 | **0.017** | -0.9 | 1.8 | 0.8 | 1.4 | 0.9 | 0.9 | -0.6 | 0.5 | -0.6 | -0.4 | -1.1 | -0.2 | -0.2 | -0.2 | -1.03 | 0.6 | -0.7 | 0.7 | -0.2 | 0.9 |
| **fnr.2 / crp.2** | Global transcriptional regulator | 0.88 | 0.0005 | -1.41 | 0.42 | -0.719 | **0.017** | -0.9 | 1.8 | 0.8 | 1.4 | 0.9 | 0.9 | -0.6 | 0.5 | -0.6 | -0.4 | -1.1 | -0.2 | -0.2 | -0.2 | -1.03 | 0.6 | -0.7 | 0.7 | -0.2 | 0.9 |
| **norV.2 / norV.1** | Flavohemoprotein NOreductase | 0.88 | 0.0005 | -1.41 | 0.42 | -0.719 | **0.017** | -0.9 | 1.8 | 0.8 | 1.4 | 0.9 | 0.9 | -0.6 | 0.5 | -0.6 | -0.4 | -1.1 | -0.2 | -0.2 | -0.1 | -1.0 | 0.6 | -0.7 | 0.7 | -0.2 | 0.9 |
| **bioB1,bioB2,bioD1,bioD2** | Biotin biosynthesis enzymes | 0.86 | 0.0008 | 3.62 | 1.24 | -5.22 | **0.026** | 0.9 | -0.2 | 1.3 | 1.4 | 0.8 | 1.2 | -1.2 | 0.4 | -1.1 | -1.0 | -0.6 | -0.4 | 0.1 | -0.7 | -1.2 | 0.7 | -0.6 | 0.7 | 0.3 | 1.1 |
| **asrC_2** | Small RNA regulator | 0.86 | 0.0007 | 3.62 | 1.24 | -5.23 | **0.024** | 0.9 | -0.2 | 1.3 | 1.4 | 0.8 | 1.2 | -1.2 | 0.4 | -1.1 | -1.0 | -0.6 | -0.4 | 0.1 | -0.7 | -1.2 | 0.7 | -0.6 | 0.7 | 0.4 | 1.1 |
| **sigV** | Sigma factor | 0.85 | 0.0016 | 1.76 | 0.59 | -3.49 | **0.042** | -0.5 | 1.1 | 0.9 | 1.6 | 0.7 | 1.5 | -0.9 | 0.6 | -0.6 | -1.5 | -0.6 | 0.2 | -0.4 | -0.3 | -0.9 | 1.1 | -0.4 | 0.8 | -0.2 | 0.7 |
| **<u>nanA.1 / nanA.2</u>** | **<u>Sialidase /neuraminidase/ *nanI*</u>** | **0.84** | **0.0019** | **3.36** | **1.16** | **-4.93** | **0.049** | **0.5** | **-0.2** | **0.9** | **1.2** | **0.7** | **1.1** | **-1.2** | **0.4** | **1.1** | **-0.9** | **-0.6** | **-0.7** | **-0.4** | **-0.6** | **-0.9** | **0.1** | **-0.6** | **0.6** | **-0.6** | **1.2** |
| **Hmp.2, hmp.1,hmp** | Flavohemoprotein | 0.84 | 0.0012 | -1.15 | 0.36 | -1.01 | **0.033** | -0.9 | 1.8 | 0.9 | 1.7 | 0.9 | 1.1 | -0.7 | 0.4 | -0.4 | -0.5 | -0.9 | -0.3 | -0.3 | -0.2 | -1.0 | 0.4 | -0.6 | 0.7 | -0.6 | 1.0 |
| **tufA, tufA-1, tuf, tuf1, tuf2** | Elongation factor Tu | 0.81 | 0.0019 | -0.84 | 0.27 | -1.3 | **0.049** | -1.5 | 1.8 | 0.8 | 1.5 | 0.8 | 1.1 | -1.2 | 0.6 | -0.9 | -1.1 | -0.6 | -0.3 | -0.2 | -0.7 | -1.0 | 0.3 | -0.9 | 0.5 | -0.5 | 1.3 |
| **rbr3A-1 / rbr3A-2** | Rubrerythrin-like protein | 0.69 | 4.51e-12 | 1.81 | 0.30 | -3.41 | **<0.01** | -1.4 | 1.9 | 0.7 | 1.4 | 0.6 | 1.3 | -1.1 | 0.6 | -0.8 | -1.2 | -0.4 | 0.0 | -0.2 | -0.8 | -1.2 | 0.2 | -0.3 | 0.7 | -0.8 | 1.1 |
| **group_3707** | hypothetical proteins | 0.63 | 0.0013 | -0.91 | 0.28 | -1.42 | **0.036** | -1.2 | 1.7 | 0.5 | 1.1 | 0.1 | 1.6 | -0.9 | 0.5 | -0.7 | -1.1 | -0.7 | -0.5 | -0.2 | -1.2 | -1.2 | -0.2 | -0.7 | 0.3 | -0.6 | 1.4 |
| **group_4463** | hypothetical proteins | 0.61 | 0.0003 | -1.03 | 0.29 | -1.36 | **0.017** | -0.9 | 1.9 | 0.5 | 1.1 | 0.1 | 1.7 | -0.8 | 0.5 | -0.7 | -1.1 | -0.7 | -0.5 | -0.2 | -1.3 | -1.1 | -0.2 | -0.7 | 0.3 | -0.7 | 1.5 |
| **group_4633** | hypothetical proteins | 0.49 | 0.0003 | 1.74 | 0.50 | -2.86 | **0.017** | -0.2 | 1.1 | 0.4 | 0.9 | 0.6 | -0.2 | -0.5 | 0.1 | -0.6 | -0.8 | -0.4 | -0.3 | -0.1 | -0.4 | -0.9 | 0.4 | -0.5 | -0.1 | 0.1 | 2.0 |
| **group_3801** | hypothetical proteins | 0.497 | 0.0003 | 1.74 | 0.50 | -2.86 | **0.017** | -0.2 | 1.1 | 0.4 | 0.9 | 0.6 | -0.2 | -0.5 | 0.1 | -0.6 | -0.8 | -0.4 | -0.3 | -0.1 | -0.4 | -0.9 | 0.4 | -0.4 | -0.2 | 0.1 | 2.0 |
| **apbE-2 / apbE-1** | hypothetical proteins | 0.497 | 0.0003 | 1.74 | 0.50 | -2.86 | **0.017** | -0.2 | 1.1 | 0.4 | 0.9 | 0.6 | -0.2 | -0.5 | 0.1 | -0.6 | -0.8 | -0.4 | -0.3 | -0.1 | -0.4 | -0.9 | 0.4 | -0.5 | -0.2 | 0.1 | 2.0 |
| **group_2511** | hypothetical proteins | 0.497 | 0.0003 | 1.74 | 0.50 | -2.86 | **0.017** | -0.9 | 1.1 | 0.4 | 0.9 | 0.6 | -0.2 | -0.5 | 0.1 | -0.6 | -0.8 | -0.4 | -0.3 | -0.1 | -0.4 | -0.9 | 0.4 | -0.5 | -0.2 | 0.0 | 2.0 |
| **group_4388** | hypothetical proteins | 0.496 | 0.0002 | 1.77 | 0.50 | -2.87 | **0.017** | -0.2 | 1.1 | 0.4 | 0.9 | 0.6 | -0.2 | -0.6 | 0.1 | -0.6 | -0.8 | -0.4 | -0.3 | -0.1 | -0.3 | -0.9 | 0.4 | -0.5 | -0.2 | 0.1 | 2.0 |
| **group_4634** | hypothetical proteins | 0.48 | 0.0003 | 0.98 | 0.27 | -2.46 | **0.017** | -0.7 | 1.5 | 1.2 | 1.2 | 1.1 | 0.9 | -1.2 | 0.7 | -0.8 | -0.6 | -0.9 | -0.3 | 0.0 | -0.4 | -0.9 | 0.7 | -0.7 | -0.0 | -0.2 | 1.7 |
| **group_2165** | hypothetical proteins | 0.439 | 0.0015 | -0.89 | 0.28 | -1.62 | **0.041** | -1.4 | 2.3 | 0.6 | 1.4 | 0.3 | 1.7 | -0.9 | 0.5 | -1.3 | -1.1 | -0.6 | -0.5 | -0.4 | -1.1 | -0.9 | -0.1 | -0.7 | 04 | -0.7 | 1.4 |
| **group_6643** | hypothetical proteins | 0.425 | 0.0009 | -0.79 | 0.24 | -1.66 | **0.029** | -1.3 | 1.5 | 1.1 | 1.3 | 0.9 | 1.2 | -1.1 | 0.6 | -0.9 | -0.9 | -0.4 | -0.7 | -0.2 | -0.5 | -0.7 | 0.5 | -0.7 | 0.3 | -0.8 | 0.8 |
| **group_6816** | hypothetical proteins | 0.399 | 0.0003 | -0.86 | 0.25 | -1.67 | **0.017** | -1.1 | 1.7 | 0.7 | 1.5 | 0.6 | 1.3 | -1 | 0.8 | -1.0 | -0.9 | -0.6 | -0.3 | -0.4 | -0.6 | -0.9 | 0.3 | -0.3 | 1.0 | -0.8 | 0.9 |
| **group_2890** | hypothetical proteins | 0.367 | 0.0005 | 0.97 | 0.28 | -2.36 | **0.017** | -1.2 | 1.7 | 0.5 | 1.1 | 0.1 | 1.6 | -0.8 | 0.5 | -0.7 | -1.1 | -0.7 | -0.6 | -0.2 | -1.2 | -1.2 | -0.2 | -0.7 | 0.3 | -0.6 | 1.4 |
| **group_2167** | hypothetical proteins | 0.367 | 0.0005 | 0.97 | 0.28 | -2.36 | **0.017** | -1.2 | 1.7 | 0.5 | 1.1 | 0.1 | 1.6 | -0.8 | 0.5 | -0.7 | -1.1 | -0.7 | -0.6 | -0.2 | -1.2 | -1.2 | -0.2 | -0.7 | 0.3 | -0.6 | 1.4 |
| **group_1125** | hypothetical proteins | 0.367 | 0.0005 | 0.97 | 0.28 | -2.36 | **0.017** | -1.2 | 1.7 | 0.5 | 1.1 | 0.1 | 1.6 | -0.8 | 0.5 | -0.7 | -1.1 | -0.7 | -0.4 | -0.2 | -1.2 | -1.2 | -0.2 | -0.7 | 0.3 | -0.6 | 1.4 |
| **regX3-2 / regX3-2** | Two-component RR | 0.366 | 0.0001 | 1.17 | 0.31 | -2.4 | **0.017** | -2.0 | 1.8 | 1.1 | 0.9 | 0.5 | 1.6 | -1.5 | 0.7 | -0.8 | -1.0 | -0.2 | -0.4 | -0.1 | -0.5 | -0.9 | 0.” | -0.6 | 0.9 | -0.§ | 0.8 |
| **group_8288** | hypothetical proteins | 0.366 | 0.0001 | 1.17 | 0.31 | -2.4 | **0.017** | -2.0 | 1.8 | 1.1 | 0.9 | 0.5 | 1.6 | -1.5 | 0.7 | -0.8 | -1.0 | -0.2 | -0.4 | -0.1 | -0.5 | -0.9 | 0.2 | -0.6 | 0.9 | -0.5 | 0.8 |

| | | | | | | | | | | | | | | | | | | | | | | | | | | | |
|---|---|---|---|---|---|---|---|---|---|---|---|---|---|---|---|---|---|---|---|---|---|---|---|---|---|---|---|
| **group_2219** | hypothetical proteins | 0.366 | 0.0001 | 1.17 | 0.31 | -2.4 | **0.017** | -2.0 | 1.8 | 1.1 | 0.9 | 0.5 | 1.6 | -1.5 | 0.7 | -0.8 | -1.0 | -0.2 | -0.4 | -0.1 | -0.5 | -0.9 | 0. | -0.6 | 0.9 | -0.5 | 0.8 |
| **group_785** | hypothetical proteins | 0.366 | 0.0001 | 1.17 | 0.31 | -2.4 | **0.017** | -2.0 | 1.8 | 1.1 | 0.9 | 0.5 | 1.6 | -1.5 | 0.7 | -0.8 | -1.0 | -0.2 | -0.4 | -0.1 | -0.5 | -0.9 | 0.2 | -0.6 | 0.9 | -0.5 | 0.8 |
| **sasA.1, sasA.2, sasA.5** | Sensor histidine kinase | 0.366 | 0.0001 | 1.17 | 0.31 | -2.4 | **0.017** | -2.0 | 1.8 | 1.1 | 0.9 | 0.5 | 1.6 | -1.5 | 0.7 | -0.8 | -1.0 | -0.2 | -0.4 | -0.1 | -0.5 | -0.9 | 0.2 | -0.6 | 0.9 | -0.5 | 0.8 |
| **group_2889** | hypothetical proteins | 0.365 | 0.0003 | 0.99 | 0.28 | -2.36 | **0.017** | -1.1 | 1.8 | 0.5 | 1.1 | 0.2 | 1.6 | -0.9 | 0.5 | -0.7 | -1.1 | -0.7 | -0.6 | -0.2 | -1.2 | -1.é | -0.2 | -0.7 | 0.” | -0.§ | 1.4 |
| **group_6859** | hypothetical proteins | 0.35 | 0.0011 | -0.81 | 0.25 | -1.72 | **0.033** | -1.1 | 1.6 | 0.8 | 1.5 | 0.6 | 1.2 | -1.1 | 0.6 | -1.1 | -0.8 | -0.6 | -0.6 | -0.2 | -0.6 | -0.! | 0.2 | -0.4 | 0.9 | -0.8 | 1.1 |
| **group_1500** | hypothetical proteins | 0.3 | 0.0003 | -0.97 | 0.27 | -1.72 | **0.017** | -0.9 | 1.6 | 0.8 | 1.5 | 0.6 | 1.5 | -1.3 | 0.34 | -0.5 | -1.2 | -0.6 | -0.2 | -0.3 | -1.2 | -1.3 | 0.3 | -0.3 | 0.§ | -0.5 | 1.0 |

Genes significantly associated with extra-intestinal isolates were identified using Pyseer (v1.3.9) under a fixed-effects model correcting for population structure with the first 20 dimensions of the MDS of the pairwise p-distance matrix.
Analyses were restricted to genes with MAF >0.25 and <0.95 to ensure >80 power to detect associations with odds ratios >2.
Each row corresponds to a gene (variant), with functional annotation where available, allele frequency (af), and association statistics. Reported values are regression coefficients (β) with standard errors, likelihood ratio test (LRT) p-values, and Benjamini-Hochberg adjusted p-values (FDR).

**Table S6**: Distribution of the resistance genes according to phylogroups, Toxinotypes, and sources

| | **Total** | **Phylogroup** | | | | | | **Toxinotype** | | | | | | | | **Source** | | | | | **Human strains: intestinal origin** | | | **Extra-intestinal strains: BWH origin** | | |
|---|---|---|---|---|---|---|---|---|---|---|---|---|---|---|---|---|---|---|---|---|---|---|---|---|---|---|
| | | **I** | **II** | **III** | **IV** | **V** | **p value** | **A** | **F** | **C** | **B** | **D** | **G** | **E** | **p value** | **human** | **animal** | **food** | **environment** | **p value** | **Intestinal** | **Extra-intestinal** | **p value** | **BWH_0** | **BWH_1** | **p value** |
| | **2231** | **202** | **106** | **1912** | **45** | **56** | | **1737** | **412** | **18** | **9** | **56** | **71** | **18** | | **1203** | **844** | **171** | **103** | | **752** | **168** | | **90** | **78** | |
| **Resistance genes** | | | | | | | | | | | | | | | | | | | | | | | | | | |
| **Tetracyclines** | | | | | | | | | | | | | | | | | | | | | | | | | | |
| ***tetA(P)*** | **1810 (78)** | 125 (61.9) | 79 (74.5) | **1537 (80.4)** | 26 (57.8) | 43 (76.8) | <0.0001 | **1447 (83.3)** | 287 (69.7) | 8 (44.4) | 0 | 16 (28.6) | 39 (54.9) | 13 (72.2) | <0.0001 | **984 (81.8)** | **637 (75.5)** | 119 (69.6) | 70 (68) | <0.0001 | 617 (82) | 139 (82.7) | | 81 (90) | 58 (74.4) | |
| ***tetB(P)*** | **1126 (48.5)** | 29 (14.4) | 29 (27.4) | **1017 (53.2)** | 23 (51.1) | 28 (50) | <0.0001 | **1021 (58.8)** | 46 (11.2) | 3 (16.7) | 0 | 14 (25) | 33 (46.5) | 9 (50) | <0.0001 | **588 (48.9)** | **454 (53.8)** | 50 (29.2) | 34 (33) | <0.0001 | 318 (42.3) | 107 (63.7) | <0.0001 | 63 (70) | 44 (56.4) | |
| ***tet(M)*** | **9 (0.4)** | 0 | 0 | 9 (0.5) | 0 | 0 | | 7 (0.4) | 0 | 0 | 0 | 1 (1.8) | 1 (1.4) | 0 | | 0 | 5 (0.6) | 2 (1.2) | 2 (1.9) | 0.0026 | 0 | 0 | | 0 | 0 | |
| ***tet(44)*** | **162 (7)** | 0 | 0 | **160 (8.4)** | 0 | 2 (3.6) | <0.0001 | **158 (9.1)** | 0 | 4 (22.2) | 0 | 0 | 0 | 0 | <0.0001 | **49 (4.1)** | 93 (11.1) | 3 (1.8) | 17 (16.5) | <0.0001 | 15 (2) | 6 (3.6) | | 4 (4.4) | 2 (2.6) | |
| ***tet(O)*** | **2 (0.1)** | 0 | 0 | 2 (0.1) | 0 | 0 | | 2 (0.1) | 0 | 0 | 0 | 0 | 0 | 0 | | 0 | 2 (0.2) | 0 | 0 | | 0 | 0 | | 0 | 0 | |
| **Macrolides** | | | | | | | | | | | | | | | | | | | | | | | | | | |
| ***erm(B)*** | **37 (1.6)** | 0 | 0 | 36 (1.9) | 1 (2.2) | 0 | | 34 (2) | 0 | 1 (5.6) | 0 | 2 (3.6) | 0 | 0 | 0.0196 | 17 (1.4) | 20 (2.4) | 0 | 0 | | 2 (0.3) | 1 (0.6) | | 0 | 1 (1.3) | |
| ***erm(Q)*** | **498 (21.5)** | 12 (5.9) | 4 (3.8) | **458 (24)** | 6 (13.3) | **18 (32.1)** | <0.0001 | **483 (27.8)** | 12 (2.9) | 0 | 0 | 3 (5.4) | 0 | 0 | <0.0001 | **244 (20.3)** | **227 (26.9)** | 12 (7) | 15 (14.6) | <0.0001 | 134 (18.2) | 14 (8.3) | 0.0235 | 7 (7.8) | 7 (9) | |
| ***erm(T)*** | **11 (0.5)** | 0 | 0 | 11 (0.6) | 0 | 0 | | 5 (0.3) | 0 | 0 | 0 | 0 | **6 (8.5)** | 0 | <0.0001 | 1 (0.1) | 7 (0.8) | 0 | 3 (2.9) | 0.0045 | 1 (0.1) | 0 | | 0 | 0 | |
| ***erm(G)*** | **8 (0.3)** | 0 | 0 | 8 (0.4) | 0 | 0 | | 8 (0.5) | 0 | 0 | 0 | 0 | 0 | 0 | | 1 (0.1) | 7 (0.8) | 0 | 0 | | 1 (0.1) | 0 | | 0 | 0 | |
| ***lnuC*** | **5 (0.2)** | 0 | 0 | 5 (0.3) | 0 | 0 | | 5 (0.3) | 0 | 0 | 0 | 0 | 0 | 0 | | 0 | 5 (0.6) | 0 | 0 | | 0 | 0 | | 0 | 0 | |
| ***lnu(P)*** | **250 (10.8)** | 2 (1) | 0 | **240 (12.6)** | 1 (2.2) | **7 (12.5)** | <0.0001 | **240 (13.8)** | 0 | 0 | 0 | 3 (5.4) | 7 (9.9) | 0 | <0.0001 | 94 (7.8) | **132 (15.7)** | 12 (7) | 12 (11.7) | <0.0001 | 40 (5.3) | 7 (4.2) | | 3 (3.3) | 4 (5.1) | |
| ***mef(A)*** | **4 (0.2)** | 0 | 0 | 4 (0.2) | 0 | 0 | | 2 (0.1) | 0 | 0 | 0 | 0 | 2 (2.8) | 0 | | 0 | 4 (0.5) | 0 | 0 | | 0 | 0 | | 0 | 0 | |
| ***optrA*** | **15 (0.6)** | 0 | 0 | 15 (0.8) | 0 | 0 | | 13 (0.7) | 0 | 0 | 0 | 2 (3.6) | 0 | 0 | | 4 (0.3) | **11 (1.3)** | 0 | **0** | | 0 | 0 | | 0 | 0 | |
| **Aminoglycosides** | | | | | | | | | | | | | | | | | | | | | | | | | | |
| ***aac(6')-Ie/aph(2'')-Ia*** | **78 (3.4)** | 0 | 0 | **77 (4)** | 1 (2.2) | 0 | 0.0019 | **74 (4.3)** | 0 | 1 (5.6) | 0 | **3 (5.4)** | 0 | 0 | <0.0001 | 31 (2.6) | **46 (5.5)** | 1 (0.6) | 0 | 0.0004 | 4 (0.5) | 3 (1.8) | | 1 (1.1) | 2 (2.6) | |
| ***aad9*** | **13 (0.6)** | 0 | 0 | 13 (0.7) | 0 | 0 | | 13 (0.7) | 0 | 0 | 0 | 0 | 0 | 0 | | 1 (0.1) | **11 (1.3)** | 0 | 1 (1) | 0.0053 | 0 | 0 | | 0 | 0 | |
| ***aadE*** | **3 (0.1)** | 0 | 0 | 3 (0.2) | 0 | 0 | | 2 (0.1) | 0 | 1 (5.6) | 0 | 0 | 0 | 0 | | 0 | 2 (0.2) | 0 | 1 (1) | | 0 | 0 | | 0 | 0 | |
| ***ant(6)-Ia*** | **12 (0.5)** | 0 | 0 | 12 (0.6) | 0 | 0 | | 12 (0.7) | 0 | 0 | 0 | 0 | 0 | 0 | | 1 (0.1) | **11 (1.3)** | 0 | 0 | 0.0073 | 1 (0.1) | 0 | | 0 | 0 | |
| ***ant(6)-Ib*** | **162 (7)** | 0 | 0 | **160 (8.4)** | 0 | 2 (3.6) | <0.0001 | **158 (9.1)** | 0 | **4 (22.2)** | 0 | 0 | 0 | 0 | <0.0001 | 48 (4) | **94 (11.1)** | 3 (1.8) | **17 (16.5)** | <0.0001 | 14 (1.9) | 6 (3.6) | | 4 (4.4) | 2 (2.6) | |
| **Phenicols** | | | | | | | | | | | | | | | | | | | | | | | | | | |
| ***fexA*** | **10 (0.4)** | 10 (0.5) | 0 | 0 | 0 | 0 | | 8 (0.5) | 0 | 0 | 0 | 2 (3.6) | 0 | 0 | | 2 (0.2) | 8 (1) | 0 | 0 | | 0 | 0 | | 0 | 0 | |
| ***cfr*** | **4 (0.2)** | 4 (0.2) | 0 | 0 | 0 | 0 | | 4 (0.2) | 0 | 0 | 0 | 0 | 0 | 0 | | 0 | 4 (0.5) | 0 | 0 | | 0 | 0 | | 0 | 0 | |

Data are presented as n (%).
p-values compare frequencies across categories using $\chi^2$ or Fisher's exact tests, corrected by the Benjamini–Hochberg.

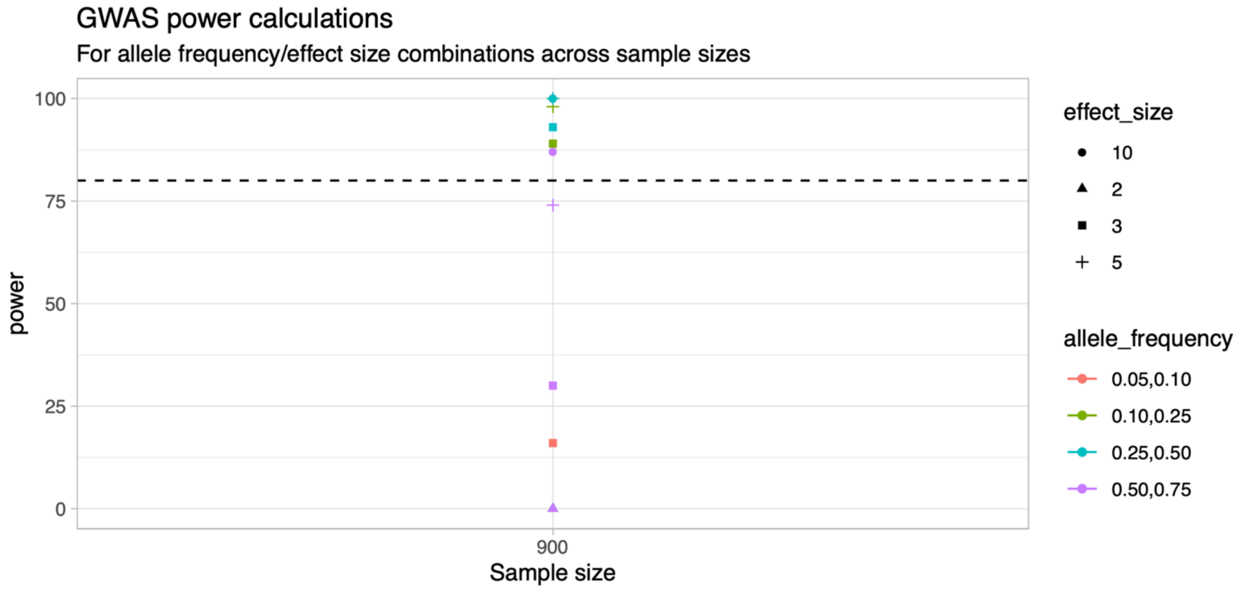


**Figure S1: GWAS power calculations by PowerBacGWAS for allele frequency and effect size combinations at the study sample size**

Power was estimated for the study sample size, accounting for the population structure using combinations of allele frequencies (0.05-0.10, 0.10-0.25, 0.25-0.50, 0.50-0.75; color-coded) and effect sizes expressed as odds ratios (OR = 2, 3, 5, 10; point shapes). The dashed horizontal line indicates the 80% power threshold. Higher power was achieved for variants with intermediate to high allele frequencies and large effect sizes (OR ≥ 3).

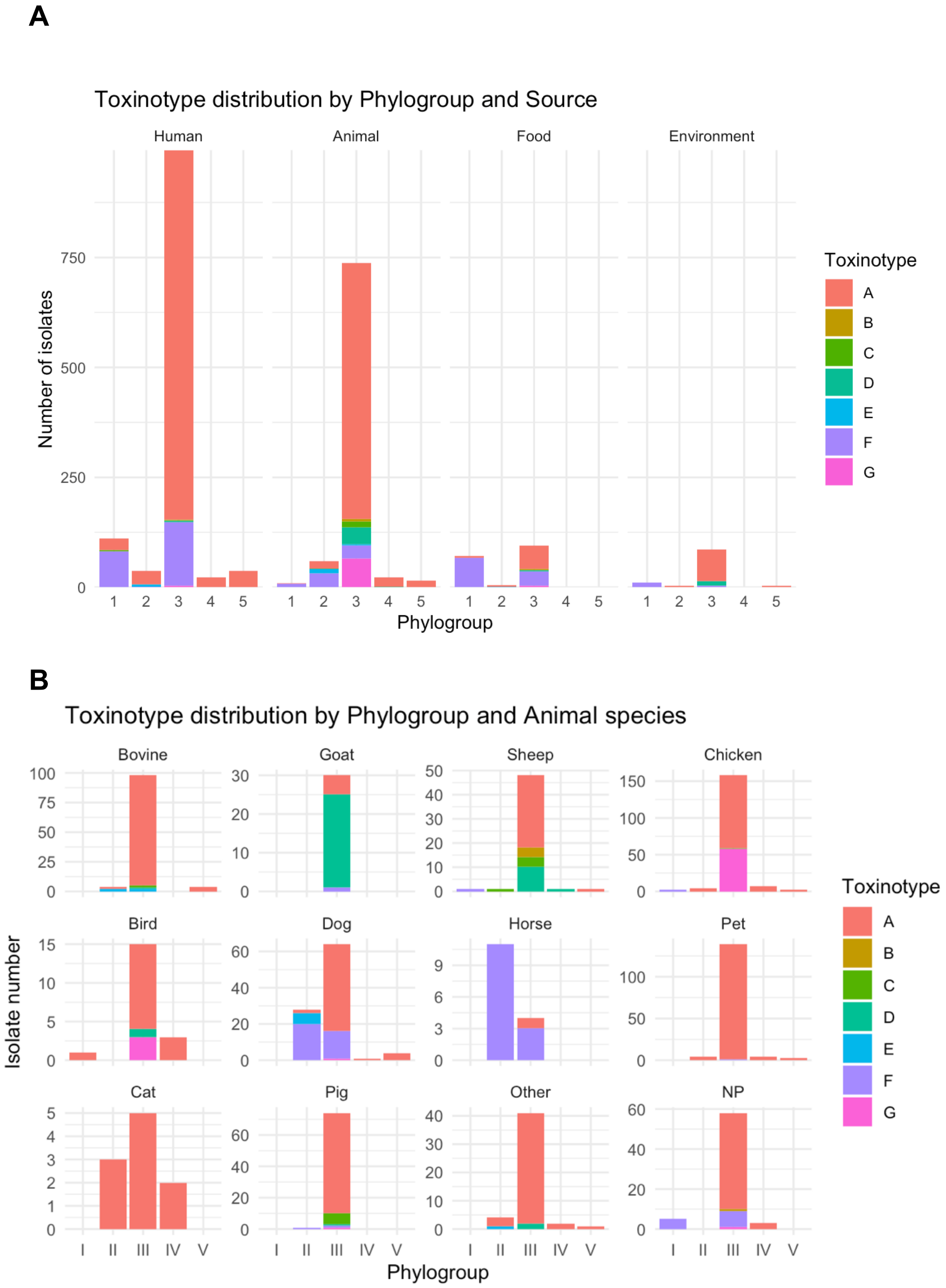


**Figure S2: Toxinotype distribution by phylogroup, source, and animal species**

**(A)** Toxinotype distribution (A-G) across phylogroups (I-V) by source (human, animal, food, environment). **(B)** Toxinotype distribution across phylogroups by animal species. Bars represent the number of isolates per toxinotype within each phylogroup-source category

A

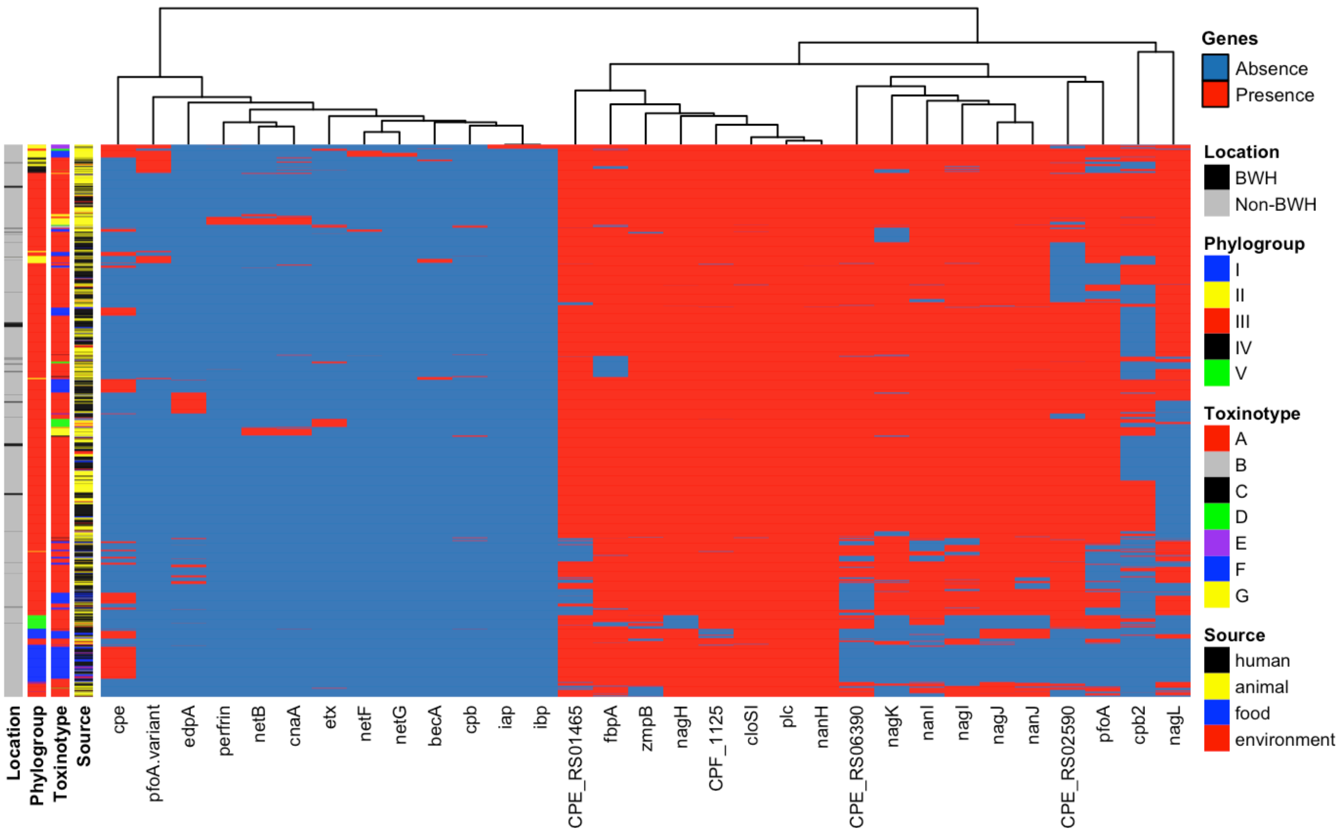
Genes
Absence
Presence
Location
BWH
Non-BWH
Phylogroup
I
II
III
IV
V
Toxinotype
A
B
C
D
E
F
G
Source
human
animal
food
environment
Location
Phylogroup
Toxinotype
Source
cpe
pfoA.variant
edpA
perfrin
netB
cnaA
etx
netF
netG
becA
cpb
iap
ibp
CPE_RS01465
fbpA
zmpB
nagH
CPF_1125
cloSI
plc
nanH
CPE_RS06390
nagK
nanI
nagI
nagJ
nanJ
CPE_RS02590
pfoA
cpb2
nagL

B

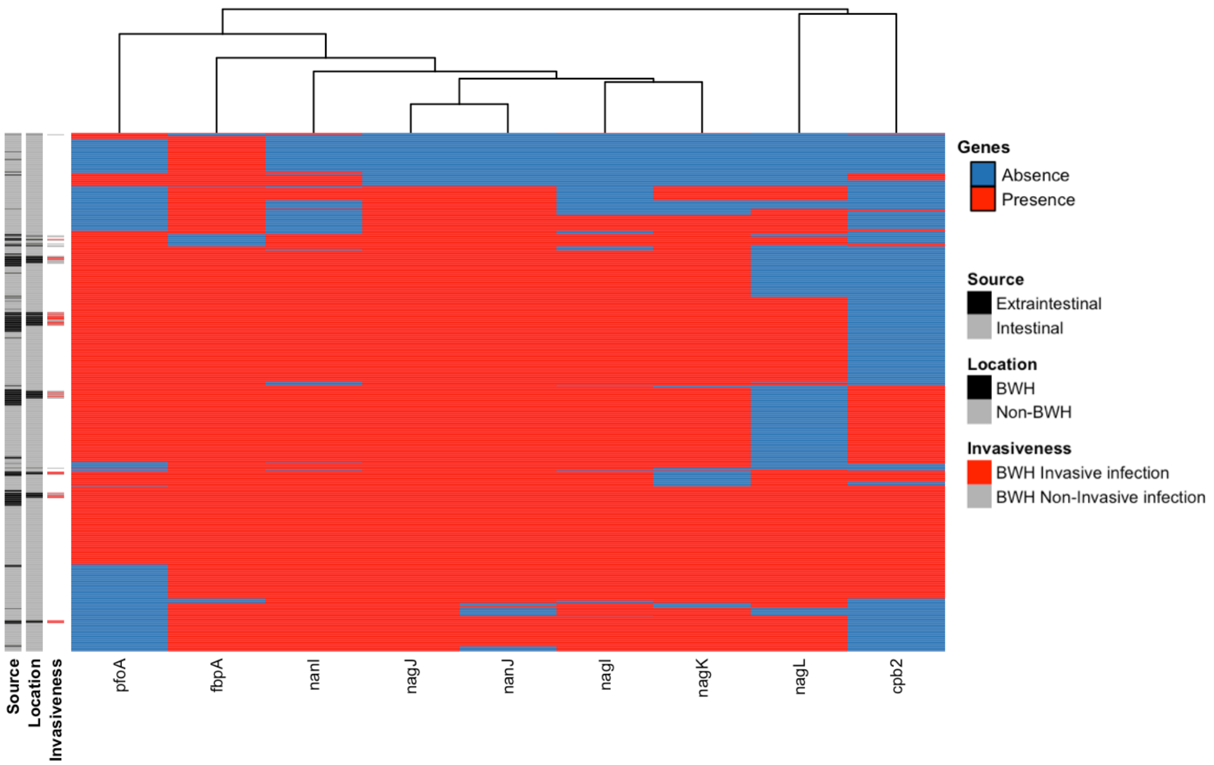
Genes
Absence
Presence
Source
Extraintestinal
Intestinal
Location
BWH
Non-BWH
Invasiveness
BWH Invasive infection
BWH Non-Invasive infection
Source
Location
Invasiveness
pfoA
fbpA
nanI
nagJ
nanJ
nagI
nagK
nagL
cpb2

**Figure S3: Virulence gene distribution by phylogroup, toxinotype, and source of isolation.**

Heatmaps show the presence (red) or absence (blue) of *C. perfringens* virulence factors across isolates. (**A**) All isolates in the dataset (n = 2,321), annotated by phylogroup (I-V), toxinotype (A-G), isolation source (human, animal, food, environment), and Brigham and Women's Hospital (BWH) location. (**B**) Human isolates (n = 920), annotated by clinical source (extraintestinal vs intestinal), BWH location, and invasiveness (BWH invasive infection, BWH non-invasive infection).
